# Rethinking Blood Eosinophils for Assessing ICS Response in COPD: A Post-Hoc Analysis from FLAME

**DOI:** 10.1101/2023.10.06.23296651

**Authors:** Alexander G. Mathioudakis, Sebastian Bate, Pradeesh Sivapalan, Jens-Ulrik Jensen, Dave Singh, Jørgen Vestbo

## Abstract

**Background:** The varied treatment response to inhaled corticosteroids (ICS) in COPD, and the increased risk of pneumonia necessitate a personalised ICS approach. This is informed by blood eosinophil count (BEC), which predicts ICS treatment response. This post-hoc analysis evaluates the ability of different BEC measurements to predict ICS treatment response. BEC measured either on or off ICS treatment, and BEC change during ICS treatment were investigated.

**Methods:** FLAME, a 52-week, double-blind RCT compared LABA/LAMA versus LABA/ICS. Corticosteroids were prohibited during a 4-week run-in period. We chose patients previously on ICS, thereby allowing pre and post run-in period BEC to represent BEC on and off ICS, respectively. In this post-hoc analysis, we revisited outcome data, exploring how the three BEC biomarkers interacted with treatment response to the ICS containing regimen.

**Results:** Our study confirms that LABA/LAMA combination is superior, or at least non-inferior, to LABA/ICS in curbing exacerbations for most FLAME participants. Lower BEC off and BEC on ICS and lack of significant BEC suppression during ICS treatment corresponded to superior response to LABA/LAMA in terms of exacerbation rate, time-to-first exacerbation, and time-to-first pneumonia. In a subgroup, including 9% of participants, BEC changed significantly during ICS treatment, and higher BEC on ICS did not predict ICS treatment response. For these patients BEC off ICS and BEC change proved more predictive.

**Conclusion:** This exploratory analysis advocates preferentially using BEC off ICS or BEC change during ICS treatment for guiding ICS treatment decisions. BEC measured on ICS is less predictive of treatment response.

**What is already known on this topic:** Blood eosinophil count (BEC) is used to guide the administration of inhaled corticosteroids (ICS) for COPD, but they may be suppressed in response to systemic or inhaled corticosteroids.

**What this study adds:** This post-hoc analysis suggests that BEC change during treatment with ICS and this change is associated with treatment response to ICS containing regimens. More specifically, BEC suppression is associated with favourable response to ICS, while unchanged or increased BEC is associated with inferior ICS treatment effect and increased risk of pneumonia. In 9% of participants, BEC changes significantly (≥200 cells/μL) during ICS treatment, and in these patients, BEC on ICS is not reliable in predicting treatment response to ICS, as it appears that some ICS responders may actually have low BEC on ICS and vice versa.

**How this study might affect research, practice or policy:** These findings highlight the potential utility of BEC change during ICS treatment as a predictive biomarker of treatment response to ICS and question the use of BEC on ICS to guide withdrawal of ICS, but need prospective validation.

## Introduction

Chronic obstructive pulmonary disease (COPD), a leading cause of death, disability, and chronic respiratory symptoms is complex and heterogeneous, thus requiring a precision medicine therapeutic approach (1). Only patients with enhanced eosinophilic inflammation in the airways appear to respond to inhaled corticosteroids (ICS) (2, 3). Blood eosinophil count (BEC), a practical surrogate marker for airway eosinophilic inflammation (2), has been shown to predict ICS treatment response in randomised controlled trials (RCTs) (4–8); higher BEC at the start of the study is associated with greater benefits for ICS treatment on exacerbations, health status and pulmonary function (2, 9). As ICS may cause pneumonia and other side effects, BECs enable a targeted ICS administration strategy in COPD patients with increased exacerbation risk (9, 10).

Systemic corticosteroids suppress BEC in COPD (3, 11), while in asthma it has been demonstrated that ICS can also suppress BEC (12, 13). A potential suppression of BEC by ICS in COPD could potentially weaken its correlation with ICS treatment response. In parallel, BEC is a responsive biomarker, as BEC suppression during treatment with corticosteroids has been associated with treatment response (11). These suggest three distinct BEC biomarkers for potential prediction of ICS treatment response: BEC measured while patients are receiving ICS (BEC on ICS); BEC off ICS; and BEC change post-ICS administration (BEC on minus BEC off ICS). Our prior post-hoc analysis of ISOLDE, a three-year RCT comparing fluticasone propionate and placebo in COPD, found BEC change to be the superior predictor (14, 15). Intriguingly, in approximately 20% of patients, ICS triggered a BEC increase, leading to a detrimental ICS effect, characterised by a surge in exacerbation rate and accelerated forced expiratory volume in 1 second (FEV_1_) decline. Moreover, higher BEC off ICS but lower BEC on ICS was associated with a slower FEV1 decline with ICS treatment. While ISOLDE was conducted in the 1990s under different care standards and less standardised exacerbation definitions, the analysis nonetheless suggests potential drawbacks in using BEC on ICS for COPD treatment decisions.

In this post-hoc analysis of the FLAME trial(16), we further evaluate whether BECs off or on ICS is better associated with treatment response to the ICS-containing regimens. We also investigated the use of BEC change as a novel biomarker of treatment response to ICS. FLAME was chosen for this analysis due to the comprehensive capture of all three BEC biomarkers in a substantial number of participants.

## Methods

This investigator-initiated, post-hoc analysis of FLAME trial was based on a prospectively designed analysis plan submitted to the study sponsor (Novartis) via Clinical Study Data Request (www.clinicalstudydatarequest.com).

### a. Overview of the FLAME trial

The FLAME trial’s methods and results have been previously published (16, 17). In brief, FLAME (NCT01782326, n=3,362), a 52-week double-blind, non-inferiority RCT, compared the effects of combining the long-acting muscarinic antagonist (LAMA) glycopyrronium and the long-acting beta-2 agonist (LABA) indacaterol, with the combination of salmeterol (LABA) and fluticasone propionate (ICS). It found the LABA/LAMA combination superior in preventing exacerbations and improving lung function and health status over the LABA/ICS combination (16). Prespecified analyses by baseline BEC indicated LABA/LAMA’s benefits as superior or similar to LABA/ICS, independent of BEC (17).

### b. Study population & BEC values

Participants underwent a 4-week run-in period while inhaled and/or systemic corticosteroids were not permitted with BEC measured before and after, the latter serving as BEC off ICS. Additionally, 56.3% of participants were already on ICS upon recruitment. In this post-hoc analysis, we included participants on ICS at baseline, who had their last dose within three days prior to the first BEC measurement (BEC on ICS). We also computed BEC change (BEC on ICS minus BEC off ICS), where a negative value indicates BEC suppression during ICS treatment. We utilised the absolute BEC values (cells/μL) in the analyses.

### c. Outcomes of interest

This post-hoc analysis investigated the correlation between three pre-specified BEC biomarkers and treatment response to the ICS-containing regimen. The primary outcomes were the rates of (i) moderate or severe and (ii) severe exacerbations. Secondary outcomes included the rate of all exacerbations, rate of exacerbations according to their treatment (only corticosteroids, only antibiotics, or both), time-to-first exacerbation, time-to-first pneumonia, change from baseline in Saint George’s Respiratory Questionnaire, in post-bronchodilator FEV_1_ and in forced vital capacity (FVC).

### d. Subgroup and sensitivity analyses

Our main analyses were reiterated for the subgroup of participants exhibiting at least a 200 cells/μL BEC change (suppression or rise). This threshold was selected by consensus among the investigators and the aim of this analysis was to exclude smaller differences in BEC that could be driven by random variability of the biomarker (18). We conducted head-to-head comparisons of LABA/LAMA and LABA/ICS in specific subgroups: patients with high BEC off ICS (≥200 cells/μL) but low BEC on ICS (<200 cells/μl) and vice versa, and those showing significant BEC suppression versus those without. In a sensitivity analysis we only incorporated participants who were administered an ICS dose within the 2 days preceding the BEC on ICS measurement.

### e. Statistical analyses

We employed appropriate statistical models to examine the interaction between treatment and BEC biomarkers. For the rate of exacerbations, we used a generalised linear model with a negative binomial distribution and included a time on treatment offset. The impact of treatment on time-to-first exacerbation, pneumonia, or death was assessed via Cox proportional hazards model. We verified the proportional hazards assumption using Kaplan-Meier curves and the Schoenfeld residuals. Furthermore, mixed-effect model repeated measures (MMRM) were used to evaluate treatment effects on change in pulmonary function and health status, focusing on the three-way interaction between treatment, time and BEC parameters.

We adjusted all analyses for age, gender, smoking status, prior exacerbation history, prior LABA or LAMA use, baseline COPD assessment test (CAT) score, and baseline FEV_1_. With less than 1% missing values for each parameter, there were no significant gaps in any covariate. All analyses were conducted in R v.3.6.3 (R core team, Vienna, Austria).

## Results

The post-hoc analysis inclusion criteria were fulfilled by 1,332 (39.6%) FLAME trial participants, split almost evenly between those receiving LABA/LAMA and LABA/ICS combinations. Baseline characteristics were similar between groups (table 1).

**Table 1.** Baseline characteristics of the participants included in this post-hoc analysis.

|  | LABA/LAMA | LABA/ICS |
| --- | --- | --- |
| Number of participants – N | 673 | 659 |
| Age – years | 64.1 (8.0) | 64.3 (7.8) |
| Female sex – no (%) | 180 (26.7%) | 192 (29.1%) |
| Current smoker – no (%) | 241 (35.8%) | 240 (36.4%) |
| COPD severity, GOLD groups – no (%) |  |  |
| COPD severity, spirometric stages – no (%) |  |  |
| GOLD 1 – Mild | 0 (0%) | 0 (0%) |
| GOLD 2 – Moderate | 206 (30.6%) | 179 (27.2%) |
| GOLD 3 – Severe | 406 (60.3%) | 422 (64.0%) |
| GOLD 4 – Very severe | 56 (8.3%) | 54 (8.2%) |
| Post-bronchodilator FEV<sub>1</sub> – % predicted | 43.7 (9.4) | 43.4 (9.3) |
| Post-bronchodilator ratio of FEV<sub>1</sub> to FVC – % | 41.1 (9.5) | 41.4 (9.8) |
| Exacerbations during the preceding year – no (%) |  |  |
| 1 | 533 (79.2%) | 510 (77.4%) |
| ≥2 | 140 (20.8%) | 149 (22.6%) |

### a. Comparison of BEC off ICS and BEC on ICS

#### Exacerbations

BEC off ICS significantly interacted with treatment effect on the rate of moderate or severe exacerbations (p for interaction <0.001). Lower BEC off ICS was related to a superior response to LABA/LAMA (intersection point: 340 cells/μL, figure 2, table S1). The model based on BEC on ICS was similar with the one based on BEC off ICS, but the interaction term did not reach statistical significance (0.069). In general, the confidence intervals for both analyses did not clearly separate due to the variability in treatment effect. Both lower BEC off and on ICS were associated with better LABA/LAMA response in terms of the time-to-first moderate or severe exacerbation (p-value <0.001, 0.008, respectively, table S2).

**Figure 1.**
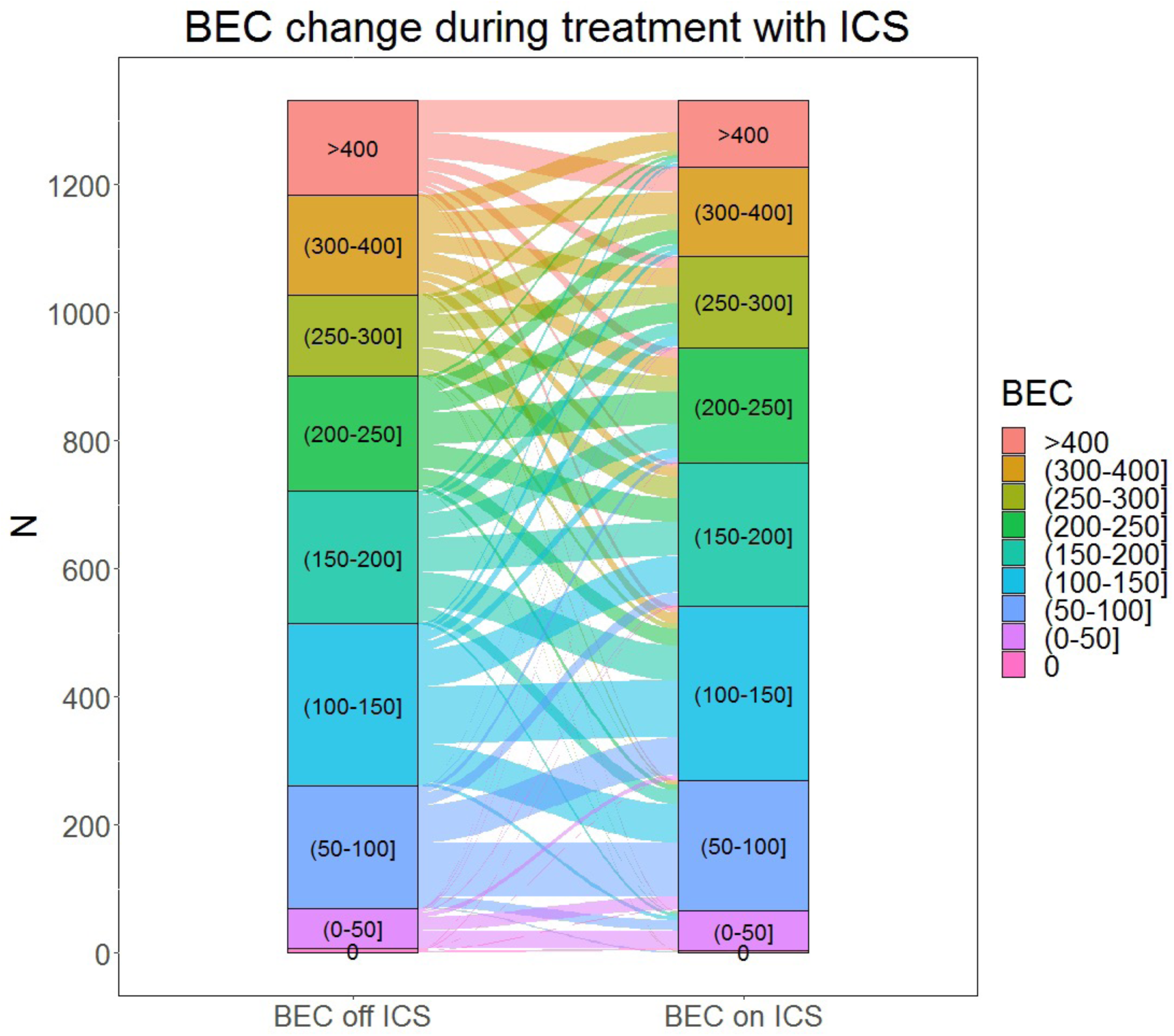
Alluvial diagram depicting BEC off ICS, BEC on ICS and BEC change during treatment with ICS among the study participants.

**Figure 2.**
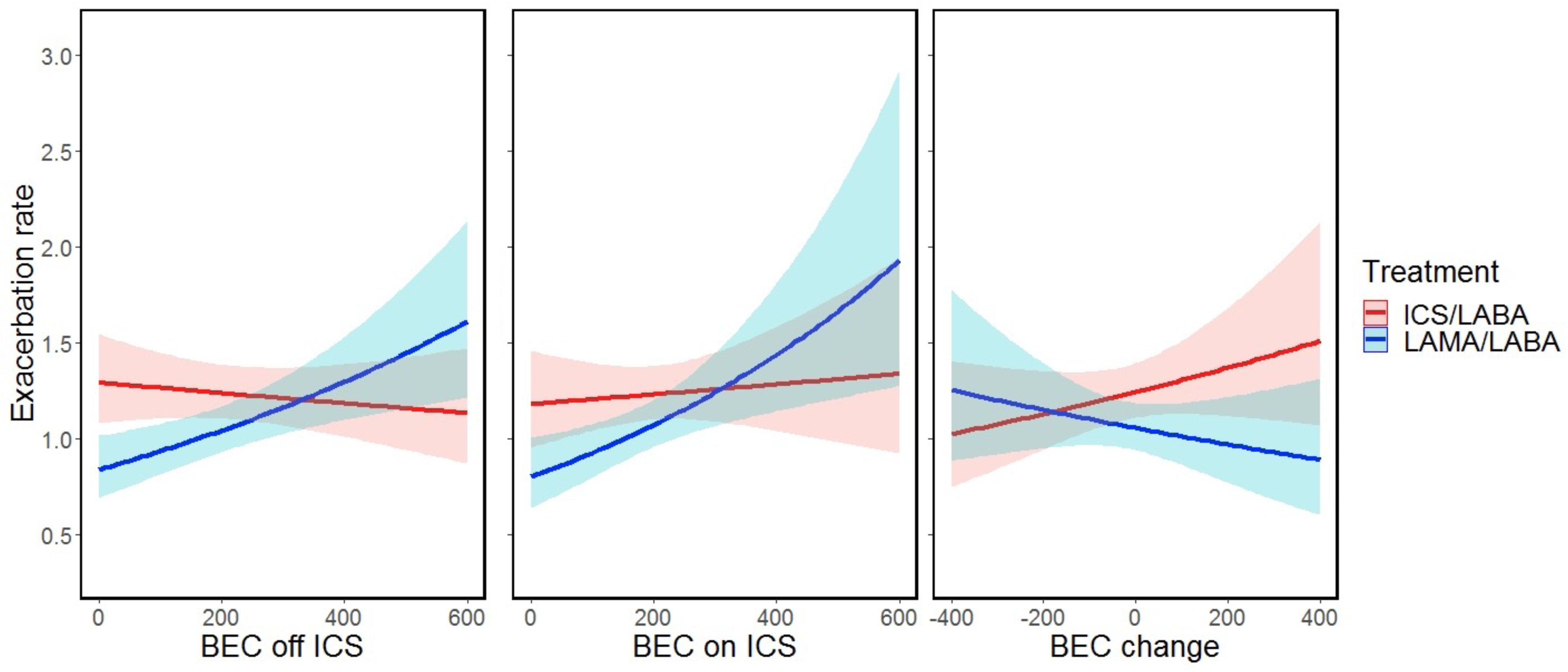
Impact of LABA/LAMA versus LABA/ICS on the frequency of **moderate or severe** exacerbations according to (a) BEC off ICS, (b) BEC on ICS, and (c) BEC change.

Neither BEC off nor on ICS demonstrated a substantial interaction with treatment effect on the rate or time-to-first severe exacerbation, perhaps due to a low event count, with only 139 participants experiencing severe exacerbations (figure S2, tables S1-2).

The association between BEC off ICS and treatment response on the rate of exacerbations of any severity (mild, moderate, or severe) did not reach statistical significance (p=0.089), but there was an association between lower BEC off ICS and favourable treatment response to LABA/LAMA on the time-to-first exacerbation of any severity (p=0.006, figure 3, tables S1-2). BEC on ICS was not associated with either of these outcomes (p= 0.886 and 0.109, respectively).

**Figure 3.**
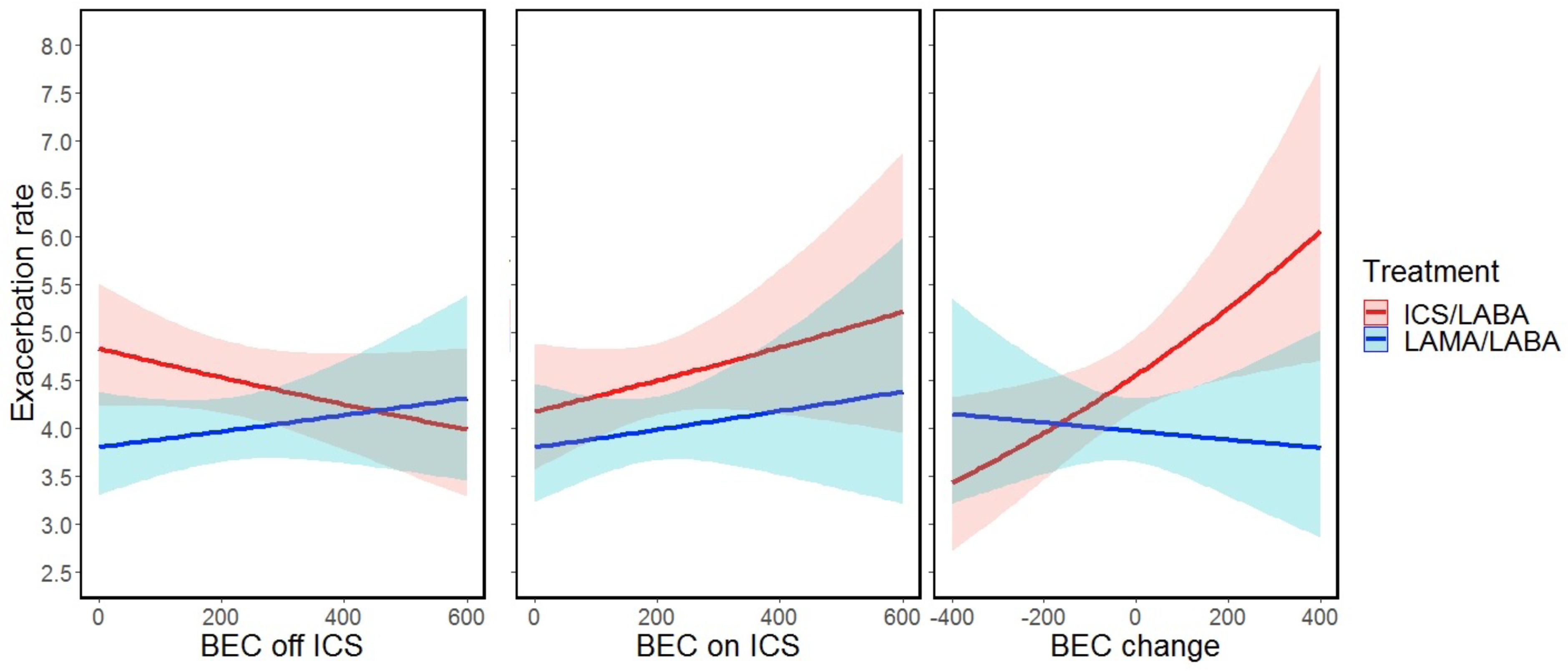
Impact of LABA/LAMA versus LABA/ICS on the frequency of **any** (mild, moderate or severe) exacerbations according to (a) BEC off ICS, (b) BEC on ICS, and (c) BEC change.

Analyses by exacerbation subtype based on treatment received (oral corticosteroids and/or antibiotics) are presented in the supplement (tables S1-2, figures S3-S5); analyses evaluating exacerbations treated with systemic corticosteroids but not antibiotics showed results that were consistent with the overall analysis of moderate or severe exacerbations.

#### Pneumonia

BEC off ICS, but not BEC on ICS, was associated with treatment response with regards to time-to-first pneumonia (p=0.041 and 0.467, respectively, table S3). According to our model, the annual risk of pneumonia among FLAME participants with a BEC off ICS of 100 cells/μL receiving LABA/ICS was 5.3%, compared to 1.8% among those receiving LABA/LAMA. The corresponding percentages for participants with BEC off ICS of 400 cells/μL were 3.9% and 3.45%, respectively.

#### Other outcomes

Neither BEC off nor on ICS were predictive of treatment response with regards to the change from baseline in pulmonary function (FEV_1_ or FVC) or health status (SGRQ).

### b. BEC change (BEC on minus BEC off ICS)

Using a BEC change threshold > 50 cells/μL to compare measurements on versus off ICS, BEC on ICS was higher in 19.5%, remained unchanged in 55.2% and decreased in 25.3% of participants. (figures 1, S1, supplement 1.1). From 738 participants with BEC off ICS <200 cells/μL, 18% showed higher values (≥200 cells/μL) while on ICS. Among 572 participants with BEC off ICS ≥200 cells/μL, 30.6% had BEC on ICS <200 cells/μL. BEC on ICS was significantly lower than BEC off ICS (median difference: -10 cells/μL, p<0.001).

### c. BEC change as a predictive therapeutic biomarker

BEC change significantly interacted with treatment effect on the rate of moderate or severe exacerbations (p=0.036), rate of exacerbations of any severity (0.041), time-to-first moderate or severe exacerbation (0.011), and time-to-first pneumonia (0.049, figure 2-3, tables S1-S3). BEC rise during ICS treatment was associated with an improved response to LABA/LAMA, while significant BEC suppression indicated an opposite trend. In our primary outcome (rate of moderate or severe exacerbations), the intersection point where LABA/LAMA appeared to be equally effective with LABA/ICS was observed at -170 cells/μL.

We grouped participants based on BEC change at the identified threshold of -170 cells/μL (figures 4-6, S6-11, S27-28). Notably, LABA/ICS was superior in reducing the rate of moderate or severe exacerbations among 104 (7.8%) participants with BEC suppression of at least 170 cells/μL (OR=0.56 [0.32, 0.98], p=0.022), while the opposite effect was observed among the 1228 (92.1%) participants with higher BEC change (> -170 cells/μL, OR=1.21 [1.03, 1.43], p<0.001). Similar trends were observed in other exacerbation outcomes and pneumonia. We also evaluated the rate of moderate or severe exacerbations using BEC change of ≤ -200, -200 to 0, and > 0 cells/μL, and observed that LABA/ICS was the superior treatment in the former group while LABA/LAMA was superior in the letter two groups (figure 6).

**Figure 4.**
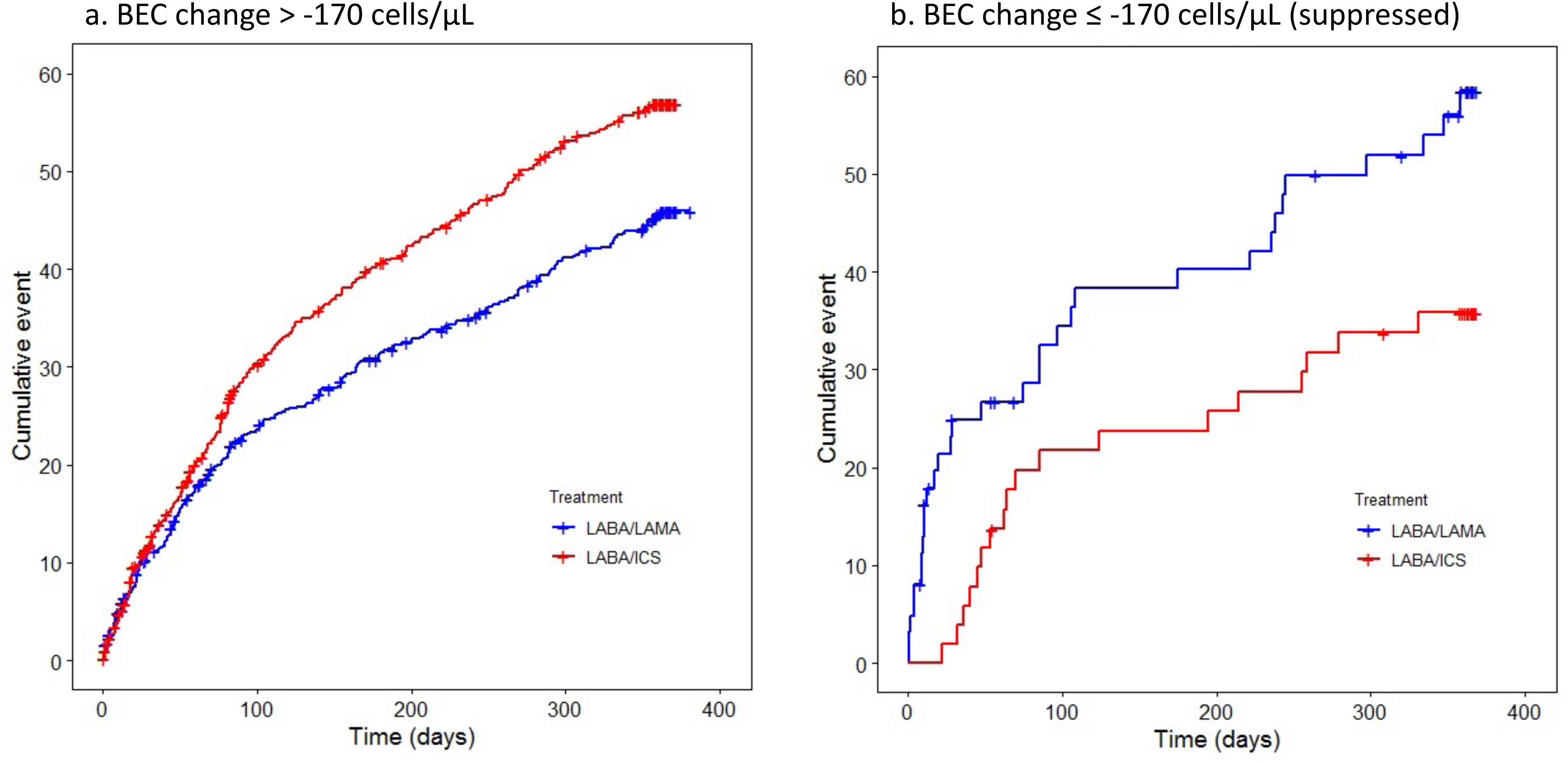
Time-to-first **moderate or severe** exacerbation stratified by treatment (LABA/LAMA vs LABA/ICS) among participants with (a) BEC change > -170 cells/μL, and (b) BEC change ≤ -170 cells/μL (significantly suppressed). The opposite treatment effects observed between patients with significantly suppressed BEC and those without should be noted.

**Figure 5.**
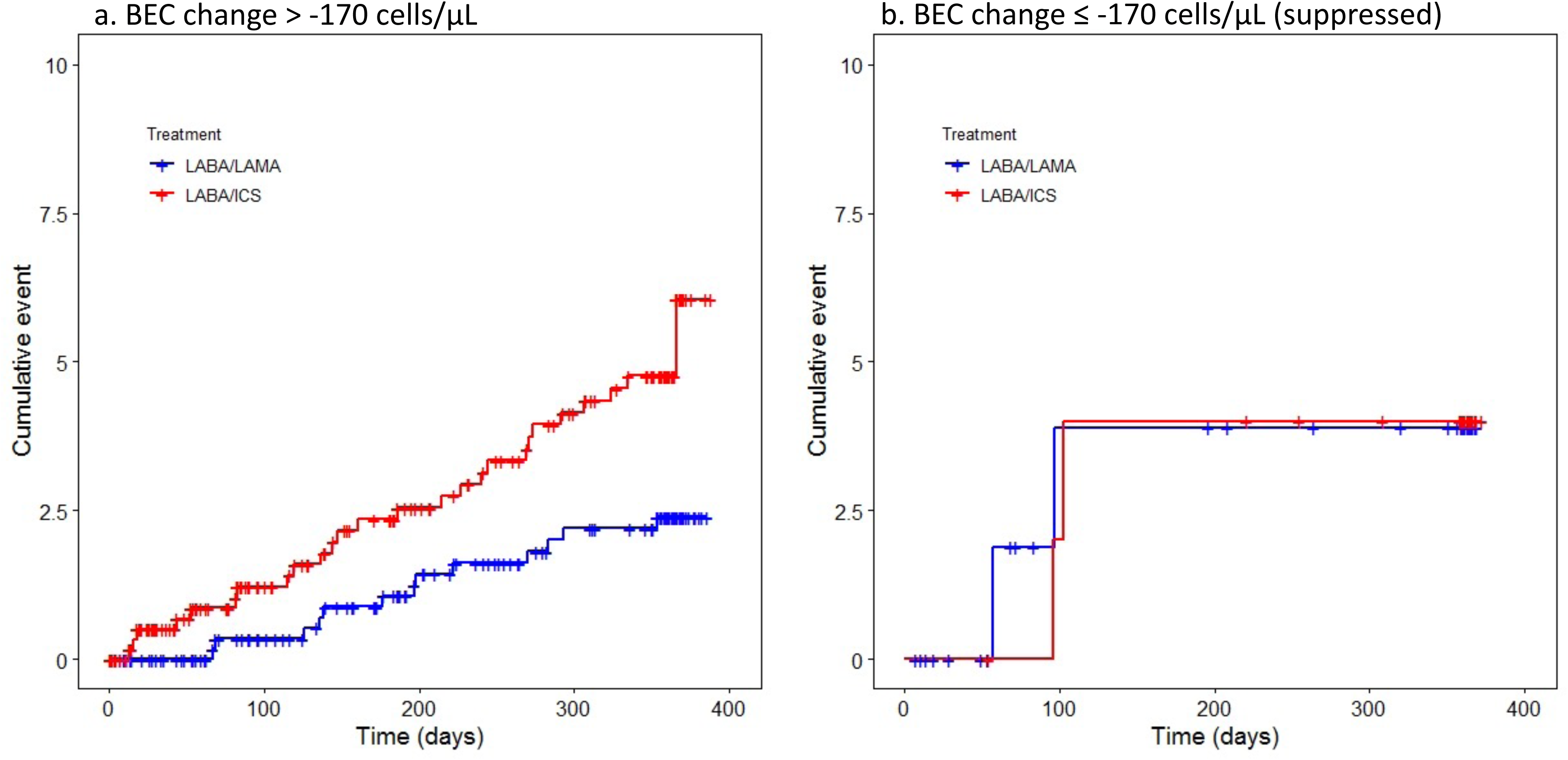
Time-to-first **episode of pneumonia** stratified by treatment (LABA/LAMA vs LABA/ICS) among participants with (a) BEC change > -170 cells/μL, and (b) BEC change ≤ -170 cells/μL (significantly suppressed). The opposite treatment effects observed between patients with significantly suppressed BEC and those without should be noted.

**Figure 6.**
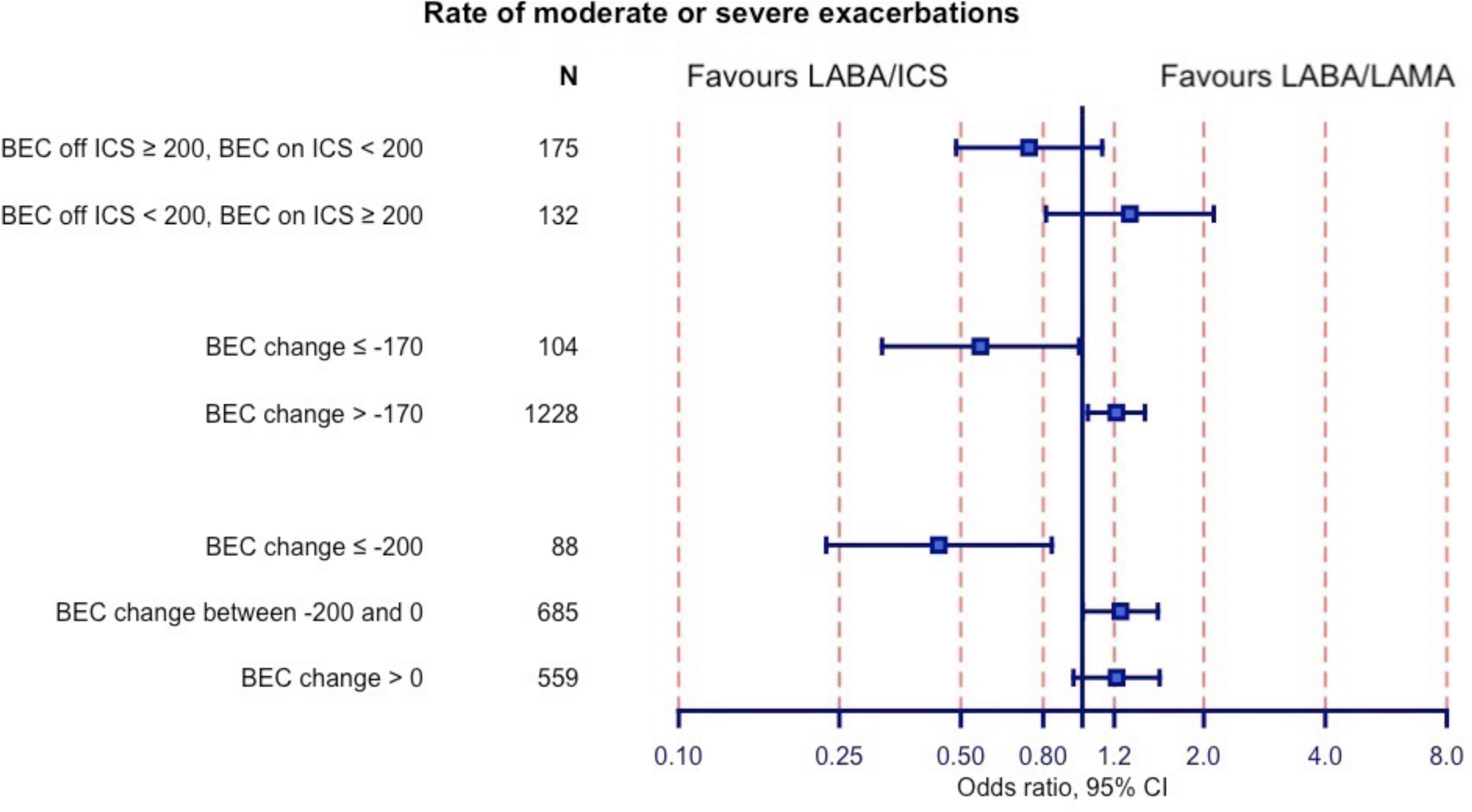
Impact of LABA/LAMA versus LABA/ICS on the rate of **moderate or severe** exacerbations in subgroup of patients with diverging BEC off versus on ICS. Odds ratio <1 favours LABA/ICS.

We evaluated the subgroup of 120 participants (9.0%) where BEC off versus on ICS differed significantly, by at least 200 cells/μL (supplement, section 3). BEC suppression was observed in 88 of these participants and BEC rise in 32. In this subgroup, higher BEC off ICS and BEC suppression both predicted improved ICS response in the rate of moderate or severe exacerbations, time-to-first moderate or severe exacerbation and time-to-first pneumonia. The interaction between treatment response and BEC on ICS was significant only for the rate of exacerbations treated with systemic corticosteroids alone. In the complementary set of participants where BEC off versus on ICS differed by <200 cells/μL, the association of both BEC off and on ICS with treatment effects on the rate of moderate or severe exacerbations was similar to the main analysis (figure S22).

Within the subgroup of 175 (13.1%) participants who had BEC off ICS ≥200 cells/μL with BEC on ICS below this threshold, there was a trend for a reduced rate of moderate or severe exacerbations favouring LABA/ICS (Odds ratio, OR=0.74, [95% confidence intervals: 0.49, 1.12], figure 6). The opposite trend was observed among 132 (9.9%) participants with BEC on ICS ≥200 cells/μL and BEC off ICS <200 cells/μL (OR=1.31 [0.81, 2.12]). Similar trends were observed in the respective subgroup analyses of the remaining exacerbation and pneumonia outcomes (supplement, section 4).

BEC change was not predictive of treatment response with regards to the change from baseline in pulmonary function or health status.

### d. Sensitivity analysis

In the subgroup of 446 (33.5%) participants who had an ICS dose within 2 days prior to BEC on ICS measurement most results were not statistically significant, likely due to the smaller sample size. Reassuringly, potential association directions were generally consistent with primary analyses (supplement, section 2).

## Discussion

This post-hoc analysis of the FLAME trial has elucidated important findings relevant to the use of blood eosinophil counts as a biomarker to personalise ICS treatment strategies in COPD. In the FLAME subgroup analysed, LABA/LAMA combination was superior or at least non-inferior to LABA/ICS in preventing exacerbations in most of the participants, consistent with the previously published results in the overall population (19). We first assessed BEC off ICS and BEC on ICS, demonstrating that higher BEC for both measurements were generally correlated with a greater prevention of moderate to severe exacerbations with ICS treatment. We performed various analyses of BEC change, as a continuous variable and using different thresholds; the FLAME dataset is unique in allowing the relationship between BEC change and treatment effect to be investigated. We show that considerable suppression of BEC may occur in some individuals which is associated with a greater ICS treatment effect. In contrast, lack of BEC suppression or an increase in BEC was a predictor of a better response to LAMA/LABA. These results highlight the potential utility of BEC change during ICS treatment as a predictive biomarker of treatment response to ICS in COPD. Furthermore, BEC off and BEC change findings highlighted excess pneumonia risk associated with ICS in patients who do not benefit from this treatment.

Several studies have demonstrated a variability of BEC in COPD(18, 20), which is greater at higher BEC. An unresolved question has been whether ICS treatment can change BEC. We show that the overall change is small (-10 cells/μL), but considerable change was observed in some individuals e.g. 9% had change ≥200 cells/μL. BEC suppression was associated with greater ICS response in different analyses, with BEC change -170 cells/μL appearing to split the population neatly into a group favouring ICS/LABA (n=104) and a group favouring LAMA/LABA treatment (n=1228). It should be noted that clinical trial populations with higher exacerbation risk (compared to FLAME) have shown a greater benefit for ICS/LABA over LAMA/LABA, so the threshold reported here will likely vary in other populations (4, 5). Nevertheless, the current analysis demonstrates that ICS can suppress BEC in COPD, and that the ICS related BEC change can be used to predict ICS responses.

BEC change suggests that BEC on ICS is not equivalent to BEC off ICS. Furthermore, the BEC change results imply that BEC on ICS may misclassify some patients with regard to ICS response prediction; e.g., higher BEC on ICS plus lower BEC off ICS seems to predict ICS non-response, but using only (higher) BEC on ICS in this individual may mistakenly predict a positive ICS response and vice versa. This can explain why BEC on ICS had less clear separation of confidence intervals for moderate to severe exacerbations compared to BEC off ICS (figure 2), and performed less well for some clinical outcomes compared to BEC off ICS e.g. frequency of any exacerbation and time to first pneumonia. The design of previous RCTs could explain why these BEC on ICS signals have been missed so far (discussion in appendix 5).

Our findings have important implications for clinical practice. First, BEC change is a new biomarker that appears to have utility and may explain ICS non-response in some individuals with higher BEC on ICS. Second, BEC on and BEC off are not identical, with BEC off appearing to offer some advantages in the prediction models performed. If BEC change is used, a relevant question is how to handle individuals without BEC change. While BEC change -200 cells/μL to 0 cells/μL appeared to favour LAMA/LABA (figure 6), there is likely a heterogeneous response within this group which is related to the absolute BEC, and a sub-analysis of individuals with < 200 cells/μL change showed that both BEC off and on ICS enabled prediction in this subgroup. Putting this all together; For the ICS naïve subject, BEC off can act as a predictor, after starting ICS the BEC change can be valuable if there is a significant change, and if not either BEC off or on ICS can be used to predict response to ICS on subjects without significant BEC change.

It should be noted that the impact of ICS on BEC may have been underestimated in this study. In FLAME, both BEC on ICS and bronchodilator reversibility were tested at the run-in visit and, in preparation of the latter, 66.5% of this post-hoc study participants did not receive ICS for three days prior to BEC measurement. While the exact timelines of BEC response to the initiation or discontinuation of ICS is uncertain, indirect data from the CORTICO-COP trial suggest effects start diminishing within 48 hours (21). Therefore, in our analysis ICS impact on BEC had likely began to diminish when BEC on ICS was measured. Indeed, in the ISOLDE post-hoc analysis, that did not have a similar limitation, an inverse association was observed between BEC on ICS and treatment response to ICS, across the whole study population (14). In general, the ISOLDE post-hoc analysis strongly supports the main findings of this analysis (details available in supplement 6).

Our analysis also demonstrated a strong association between BEC biomarkers and pneumonia risk among patients with COPD receiving ICS. The heightened pneumonia risk was primarily observed in patients unlikely to benefit from ICS. This aligns with past research indicating susceptibility to bacterial infections in non-eosinophilic COPD (22–24) and reinforces the need to target ICS use to avoid unnecessary risks. We did not establish a link between BEC variables and treatment effect on pulmonary function and health status trajectories. This could be due to the short follow-up period of one year, which might not be sufficient to evaluate treatment impact on pulmonary function decline rate. Furthermore, lack of between treatment differences are not surprising since dual bronchodilators are known for their excellent activity on these outcomes (25).

The findings from the two post-hoc analyses of the ISOLDE and FLAME trials highlight the need for a prospective study of BEC assessment timing and efficacy of ICS. They also question the use of BEC to guide withdrawal of ICS, as it appears that some ICS responders may actually have low BEC on ICS and vice versa. BEC remain a predictor of ICS response, but our data adds some complexity to the way that BEC could be interpreted.

## Data Availability

Data from the FLAME trial are available upon request through the clinical study data request platform (clinicalstudydatarequest.com. Applications are reviewed for merit by an independent review board before access is granted.

## Author Contributions

Study Conception: AGM, JV. Study design: AGM, JV. Statistical analysis: AGM, SB. Interpretation of findings: All authors. Manuscript drafting: AGM, DS, JV. Critical revision of the manuscript: All authors.

## Acknowledgement

The authors are grateful to the FLAME investigators and participants and to Novartis and Clinical Study Data Request (CSDR, a consortium of clinical study sponsors and funders that was developed to facilitate patient level data sharing, ClinicalStudyDataRequest.com) for providing access to patient level data from the FLAME trial.

