## supplement for "Rethinking Blood Eosinophils for Assessing ICS Response in COPD: A Post-Hoc Analysis from FLAME"

[Figure S27. Time-to-first **moderate or severe exacerbation.** Hazard ratio <1 favours LABA/ICS.	42](#_Toc141117438)

[Figure S28. Time-to-first episode of **pneumonia** Hazard ratio <1 favours LABA/ICS.	43](#_Toc141117439)

### 1. Main analyses: Further data and figures.

#### 1.1. BEC change after ICS administration.

Considering BEC unchanged if the difference between the two variables (BEC on and off ICS) was less than 10 cells/μL, BEC on ICS was raised in 41.9%, unchanged in 5.5% and decreased in 52.6% of participants. When changing the threshold to 50 cell/μL, then BEC on ICS was raised in 19.5%, unchanged in 55.2% and decreased in 25.3% of participants compared to BEC off ICS.

##### Figure S1: Association between BEC off ICS and BEC on ICS.

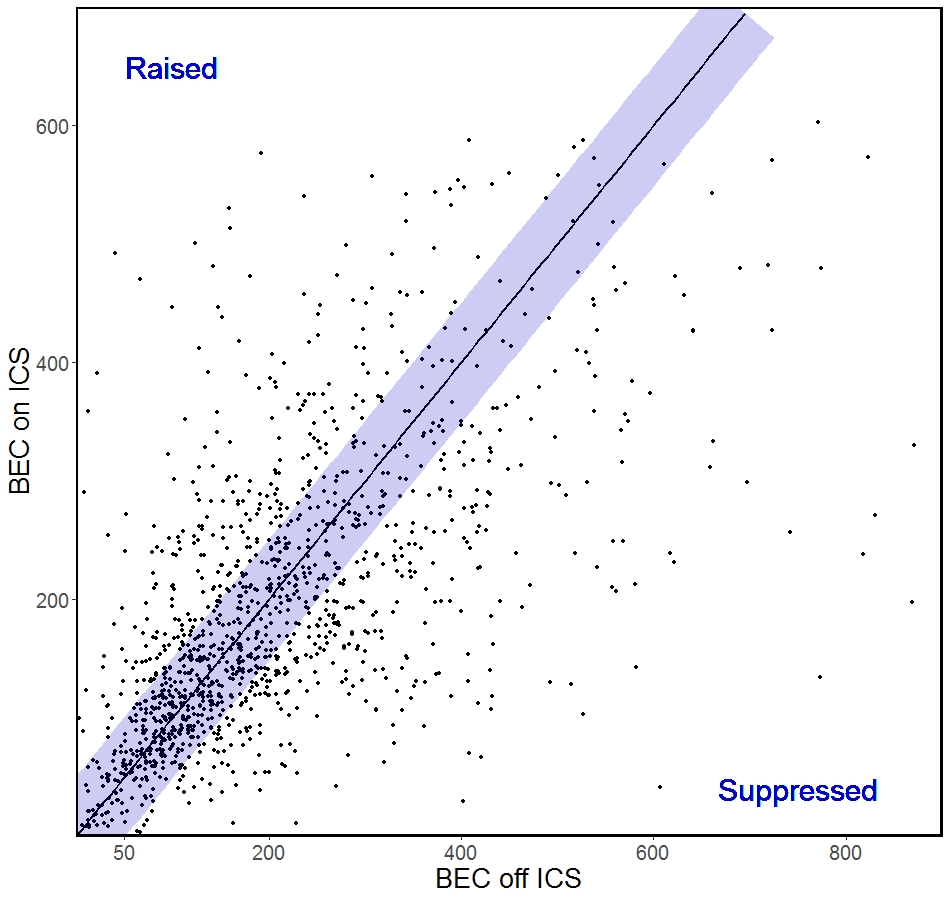

**R = 0.549**

#### 1.3. Frequency of exacerbations

##### Table S1. Impact of LABA/LAMA versus LABA/ICS on the frequency of exacerbations. Summary of the main results of the general linear regression analysis.

| Relevant  Parameters  Analysis | BEC | | ICS/LABA | | Three-way interaction | |
| --- | --- | --- | --- | --- | --- | --- |
|  | Coeff  (cells / mL) | p | Coeff  (Yes) | p | Coeff  (cells / mL) | p |
| **Rate of moderate or severe exacerbations**  BEC off ICS  BEC on ICS  BEC change | 1.189  1.300  -0.728 | **<0.001**  **0.008**  0.081 | 0.449  0.363  0.154 | **<0.001**  **0.019**  0.057 | -1.457  -1.203  1.190 | **0.001**  0.069  **0.036** |
| **Rate of severe exacerbations**  BEC off ICS  BEC on ICS  BEC change | 1.351  0.654  -1.424 | 0.076  0.578  0.121 | 0.151  0.029  0.067 | 0.621  0.938  0.736 | -0.564  0.054  0.781 | 0.574  0.973  0.515 |
| **Rate of any exacerbation**  BEC off ICS  BEC on ICS  BEC change | 0.262  0.225  -0.194 | 0.330  0.544  0.545 | 0.234  0.086  0.119 | 0.016  0.452  0.046 | -0.604  0.071  0.881 | 0.089  0.886  **0.041** |
| **Moderate or severe exacerbations treated with systemic corticosteroids and not antibiotics**  BEC off ICS  BEC on ICS  BEC change | 2.130  2.644  -1.509 | **<0.001**  0.010  0.068 | 0.770  0.733  0.206 | **0.005**  **0.028**  0.235 | -2.751  -2.867  2.274 | **0.004**  0.041  0.056 |
| **Moderate or severe exacerbations treated with antibiotics and not systemic corticosteroids**  BEC off ICS  BEC on ICS  BEC change | 0.747  -0.346  -1.171 | 0.221  0.697  0.097 | 0.392  0.109  0.162 | 0.088  0.682  0.249 | -1.249  0.063  1.705 | 0.134  0.958  0.087 |
| **Moderate or severe exacerbations treated with both antibiotics and systemic corticosteroids**  BEC off ICS  BEC on ICS  BEC change | 0.874  1.584  0.058 | 0.053  **0.015**  0.923 | 0.338  0.368  0.155 | 0.051  0.076  0.153 | -0.882  -1.145  0.398 | 0.145  0.190  0.610 |

##### Figure S2. Impact of LABA/LAMA versus LABA/ICS on the frequency of **severe** exacerbations according to (a) BEC off ICS, (b) BEC on ICS, and (c) BEC change.

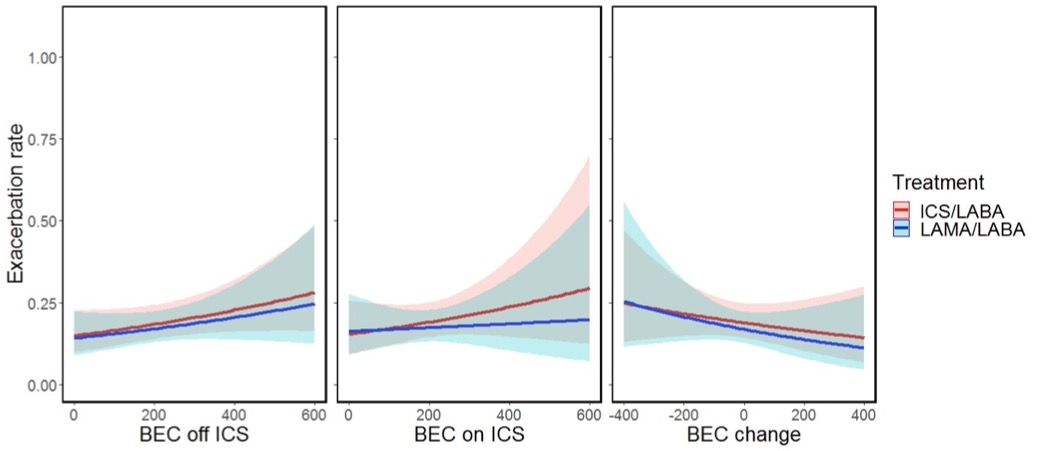

##### Figure S3. Impact of LABA/LAMA versus LABA/ICS on the frequency of **moderate or** **severe exacerbations treated with systemic corticosteroids and not with antibiotics** according to (a) BEC off ICS, (b) BEC on ICS, and (c) BEC change.

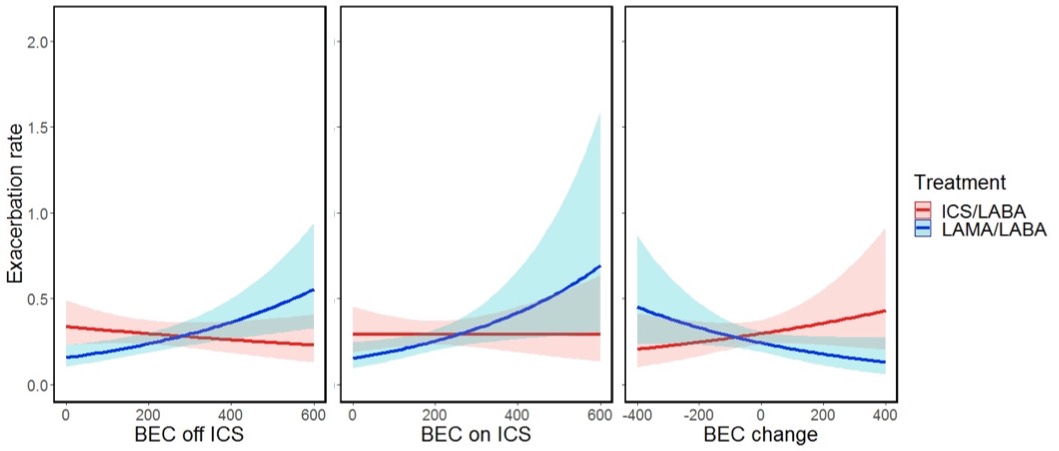

##### Figure S4. Impact of LABA/LAMA versus LABA/ICS on the frequency of **moderate or** **severe exacerbations treated with antibiotics and not with systemic corticosteroids** according to (a) BEC off ICS, (b) BEC on ICS, and (c) BEC change.

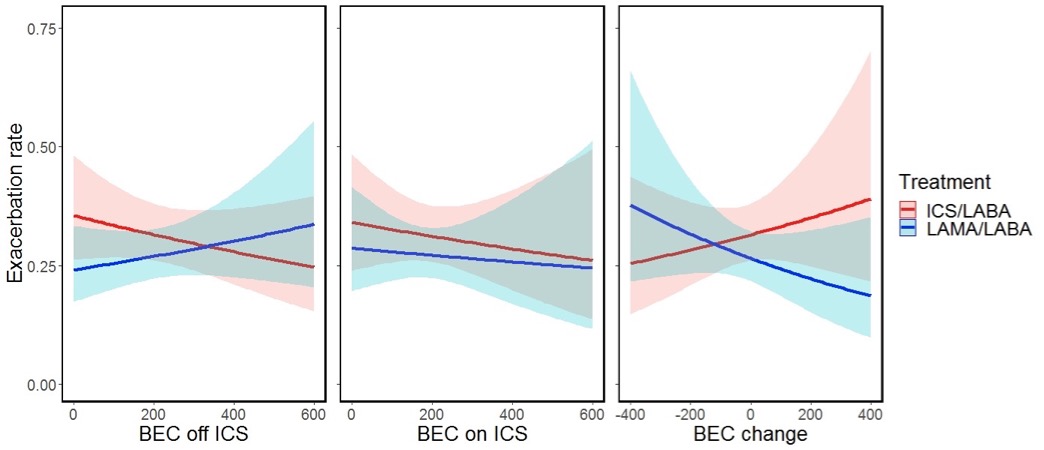

##### Figure S5. Impact of LABA/LAMA versus LABA/ICS on the frequency of **moderate or** **severe exacerbations treated with antibiotics and systemic corticosteroids** according to (a) BEC off ICS, (b) BEC on ICS, and (c) BEC change.

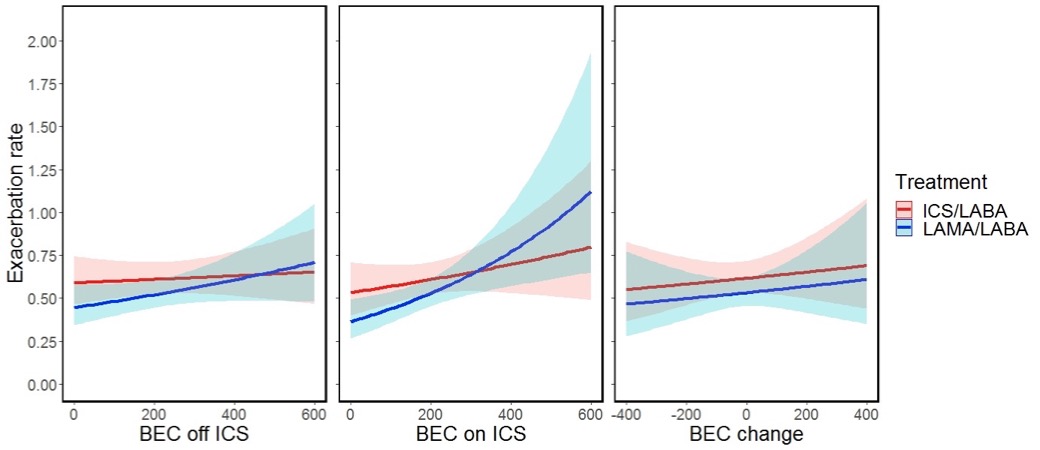

#### 1.4. Time to first exacerbation

##### Table S2: Impact of LABA/LAMA versus LABA/ICS on the time to first exacerbation. Summary of the main results of the Cox proportional hazards analysis.

| Relevant  Parameters  Analysis | BEC | | ICS/LABA | | Three-way interaction | |
| --- | --- | --- | --- | --- | --- | --- |
|  | Coeff  (cells / mL) | p | Coeff  (Yes) | p | Coeff  (cells / mL) | p |
| **Time-to-first moderate or severe exacerbation**  BEC off ICS  BEC on ICS  BEC change | 1.335  1.631  -1.020 | **<0.001**  **<0.001**  **0.015** | 0.574  0.544  0.235 | **<0.001**  **<0.001**  **0.004** | -1.702  -1.758  1.449 | **<0.001**  **0.008**  **0.011** |
| **Time-to-first severe exacerbation**  BEC off ICS  BEC on ICS  BEC change | 1.594  1.638  -1.342 | **0.011**  0.120  0.113 | 0.338  0.281  0.210 | 0.206  0.412  0.244 | -0.708  -0.514  0.698 | 0.369  0.709  0.498 |
| **Time-to-first exacerbation (any severity)**  BEC off ICS  BEC on ICS  BEC change | 0.467  0.494  -0.336 | 0.071  0.187  0.310 | 0.316  0.261  0.118 | **0.002**  **0.029**  0.060 | -0.988  -0.828  0.818 | **0.006**  0.109  0.062 |
| **Time-to-first moderate or severe exacerbation treated with systemic corticosteroids and not antibiotics**  BEC off ICS  BEC on ICS  BEC change | 1.890  2.986  -1.174 | **<0.001**  **<0.001**  0.095 | 0.620  0.738  0.194 | **0.007**  **0.011**  0.194 | -2.026  -2.837  1.483 | **0.005**  **0.015**  0.135 |
| **Time-to-first moderate or severe exacerbation treated with antibiotics and not systemic corticosteroids**  BEC off ICS  BEC on ICS  BEC change | 0.786  -0.026  -1.143 | 0.149  0.975  0.072 | 0.289  0.049  0.052 | 0.185  0.844  0.696 | -1.281  -0.203  1.702 | 0.104  0.855  0.073 |
| **Time-to-first moderate or severe exacerbation treated with both antibiotics and systemic corticosteroids**  BEC off ICS  BEC on ICS  BEC change | 0.933  1.665  -0.135 | 0.030  **0.009**  0.832 | 0.522  0.553  0.293 | **0.002**  **0.006**  **0.005** | -1.109  -1.399  0.581 | 0.057  0.098  0.470 |

##### Figure S6: Time-to-first **severe** exacerbation stratified by treatment (QVA vs SALFL) and BEC suppression by at least 170 cells/μL (-) or higher BEC change levels (+).

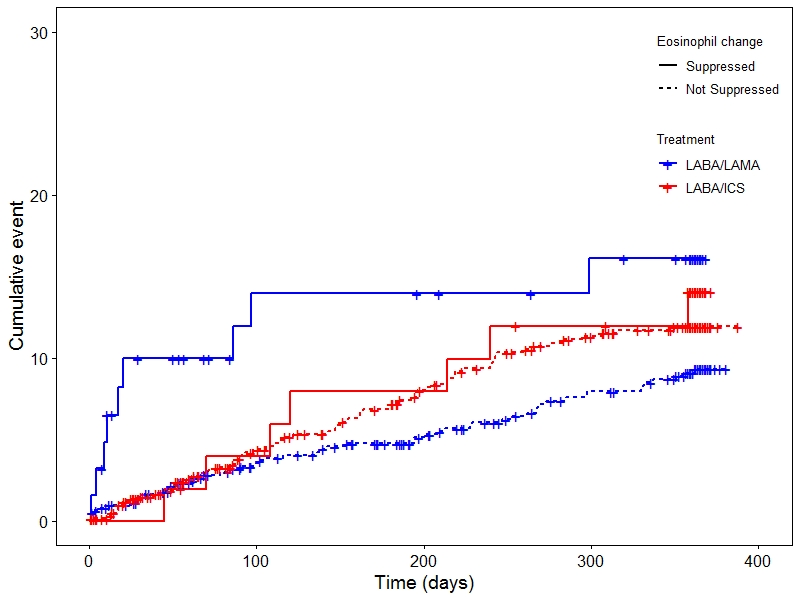

##### Figure S7: Time-to-first **exacerbation of any severity (mild, moderate or severe)** stratified by treatment (QVA vs SALFL) and BEC suppression by at least 170 cells/μL (-) or higher BEC change levels (+).

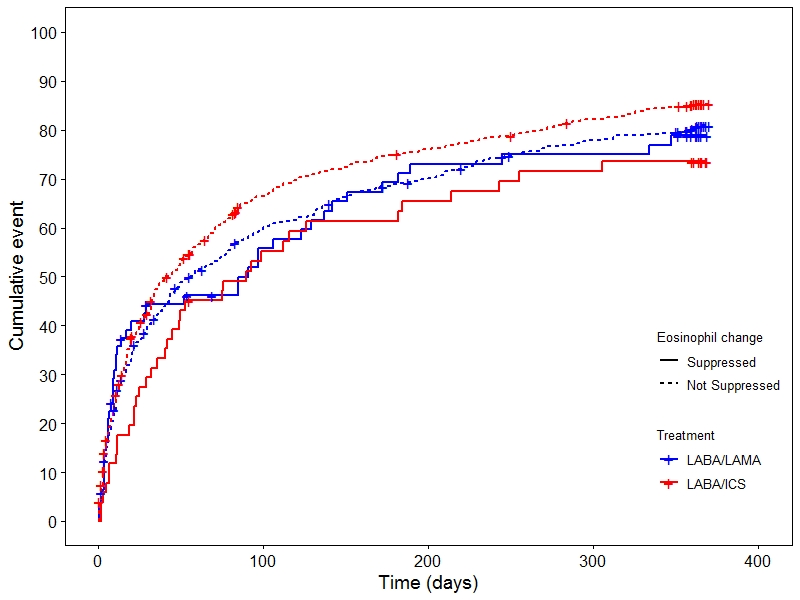

##### Figure S8: Time-to-first **moderate or severe** **exacerbation treated with systemic corticosteroids and without antibiotics** stratified by treatment (QVA vs SALFL) and BEC suppression by at least 170 cells/μL (-) or higher BEC change levels (+).

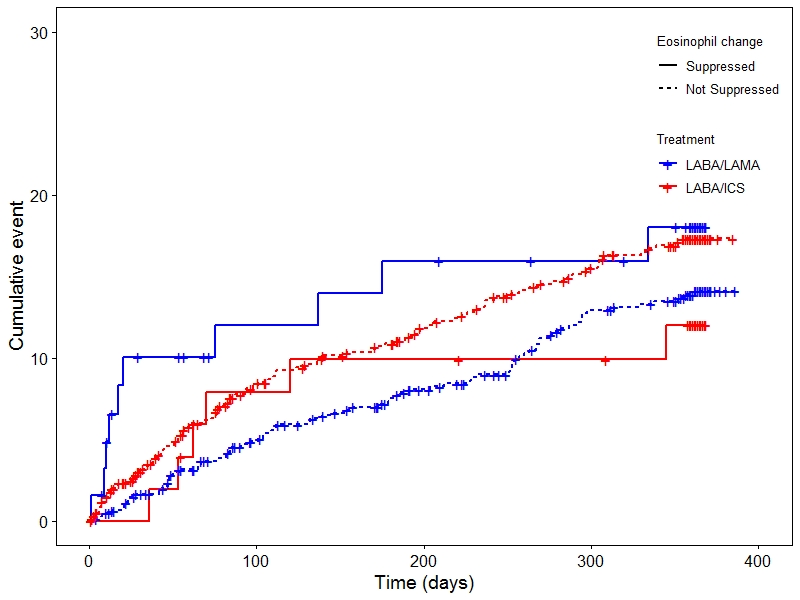

##### Figure S9: Time-to-first **moderate or severe** **exacerbation treated with antibiotics and without systemic corticosteroids** stratified by treatment (QVA vs SALFL) and BEC suppression by at least 170 cells/μL (-) or higher BEC change levels (+).

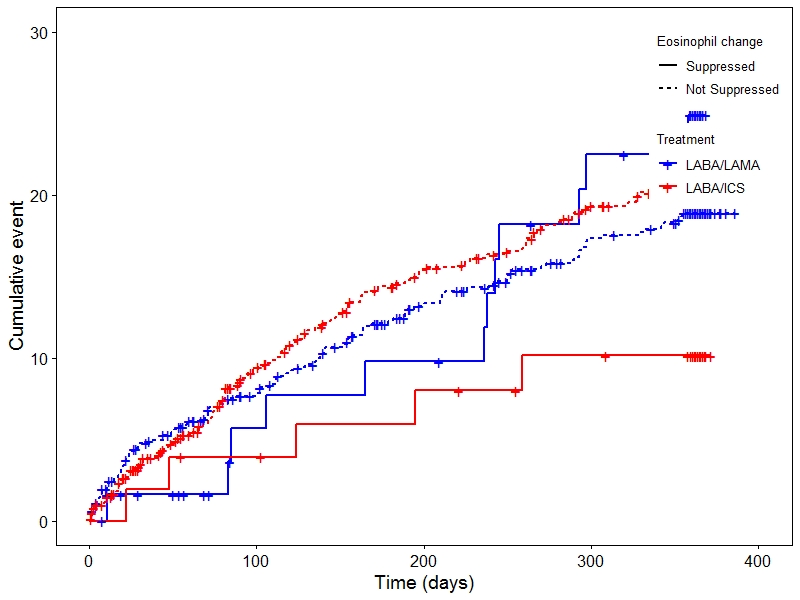

##### Figure S10: Time-to-first **moderate or severe** **exacerbation treated with systemic corticosteroids and antibiotics** stratified by treatment (QVA vs SALFL) and BEC suppression by at least 170 cells/μL (-) or higher BEC change levels (+).

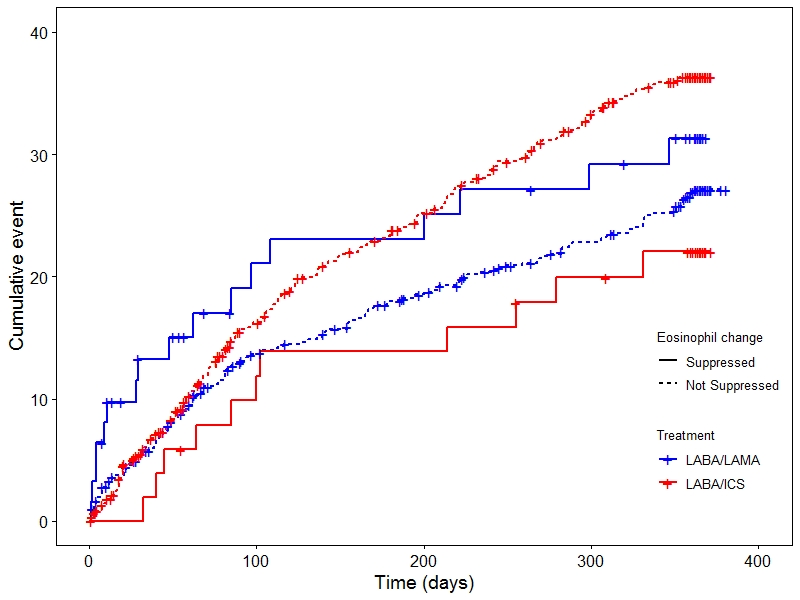

#### 1.5. Time to pneumonia

##### Table S3: Impact of LABA/LAMA versus LABA/ICS on the time to first episode of pneumonia. Summary of the main results of the Cox proportional hazards analysis.

| Relevant  Parameters  Analysis | BEC | | ICS/LABA | | Three-way interaction | |
| --- | --- | --- | --- | --- | --- | --- |
|  | Coeff  (cells / mL) | p | Coeff  (Yes) | p | Coeff  (cells / mL) | p |
| **Time to first episode of pneumonia**  BEC off ICS  BEC on ICS  BEC change | 2.409  2.044  -2.564 | **0.033**  0.347  0.074 | 1.436  1.018  0.771 | **0.006**  0.112  **0.023** | -3.428  -1.931  4.257 | **0.042**  0.467  **0.049** |

### 2. Subgroup analysis only including participants that received a dose of ICS within 2 days form the BEC on ICS measurement.

#### 2.1. BEC change after ICS administration.

##### Figure S11: Alluvial diagram depicting BEC off ICS, BEC on ICS and BEC change during treatment with ICS among the study participants that received a dose of ICS within 2 days prior the BEC on ICS measurements.

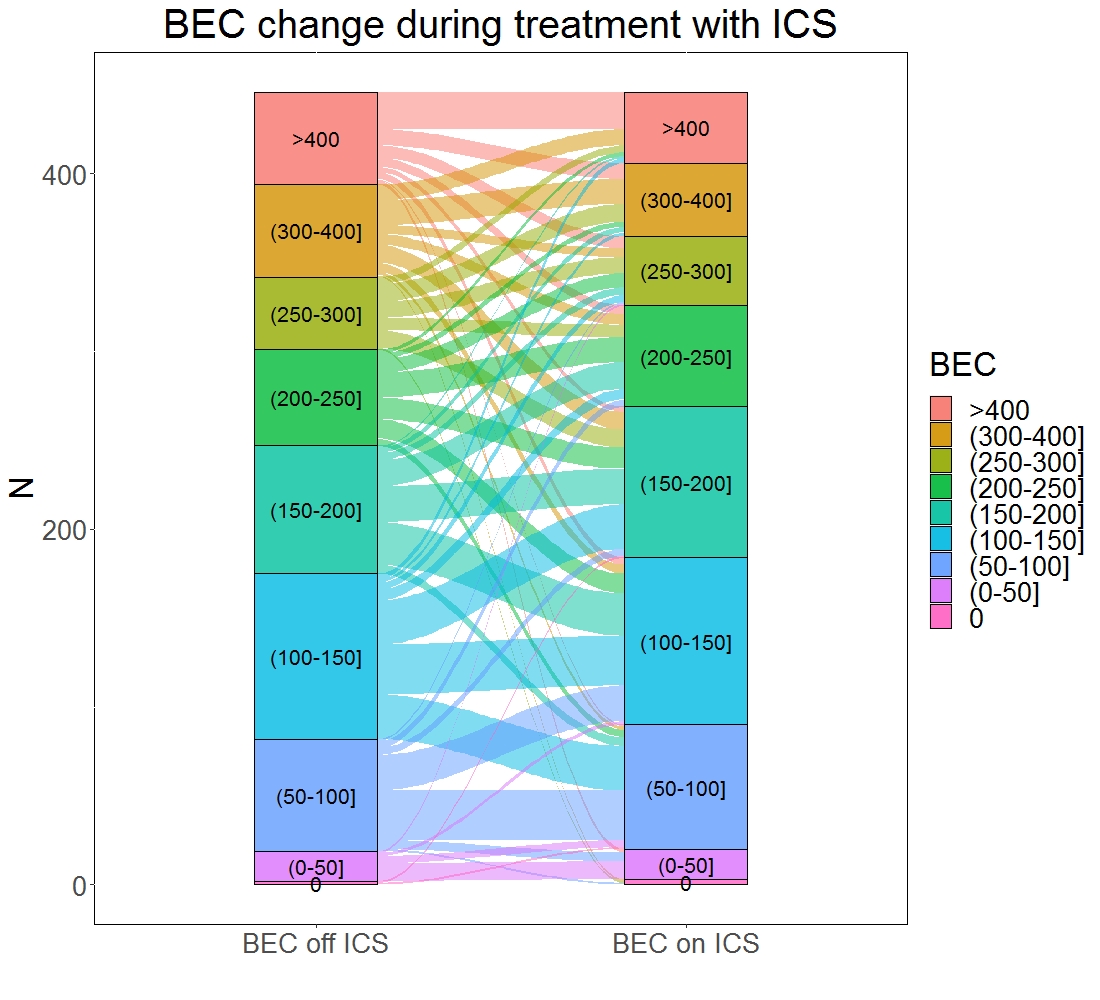

#### 2.2. Frequency of exacerbations

##### Table S4: Impact of LABA/LAMA versus LABA/ICS on the frequency of exacerbations. Subgroup analysis only including patients that received the last dose of their ICS within 2 days from the BEC on ICS measurement. Summary of the main results of the general linear regression analysis.

| Relevant  Parameters  Analysis | BEC | | ICS/LABA | | Three-way interaction | |
| --- | --- | --- | --- | --- | --- | --- |
|  | Coeff  (cells / mL) | p | Coeff  (Yes) | p | Coeff  (cells / mL) | p |
| **Rate of moderate or severe exacerbations**  BEC off ICS  BEC on ICS  BEC change | 1.240  1.835  -0.883 | **0.003**  **0.025**  0.092 | 0.262  0.292  0.035 | 0.205  0.253  0.796 | -1.082  -1.532  0.970 | 0.114  0.148  0.274 |
| **Rate of severe exacerbations**  BEC off ICS  BEC on ICS  BEC change | 1.407  1.795  -1.257 | 0.174  0.407  0.327 | 0.095  0.315  0.020 | 0.861  0.643  0.957 | -0.475  -1.791  0.219 | 0.777  0.523  0.918 |

##### Figure S12. Impact of LABA/LAMA versus LABA/ICS on the frequency of **moderate or severe exacerbations** according to (a) BEC off ICS, (b) BEC on ICS, and (c) BEC change. **Subgroup analysis only including participants that received a dose of ICS within 2 days form the BEC on ICS measurement.**

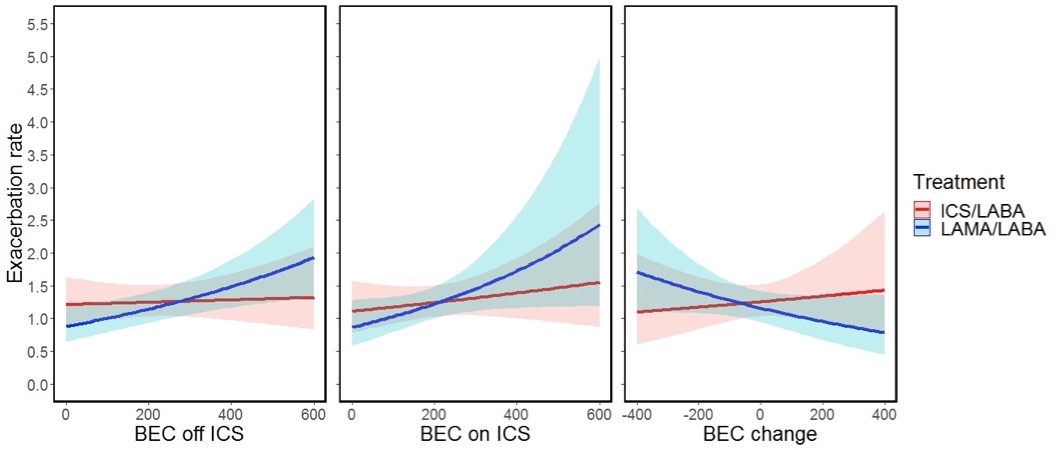

##### Figure S13. Impact of LABA/LAMA versus LABA/ICS on the frequency of **severe exacerbations** according to (a) BEC off ICS, (b) BEC on ICS, and (c) BEC change. **Subgroup analysis only including participants that received a dose of ICS within 2 days form the BEC on ICS measurement.**

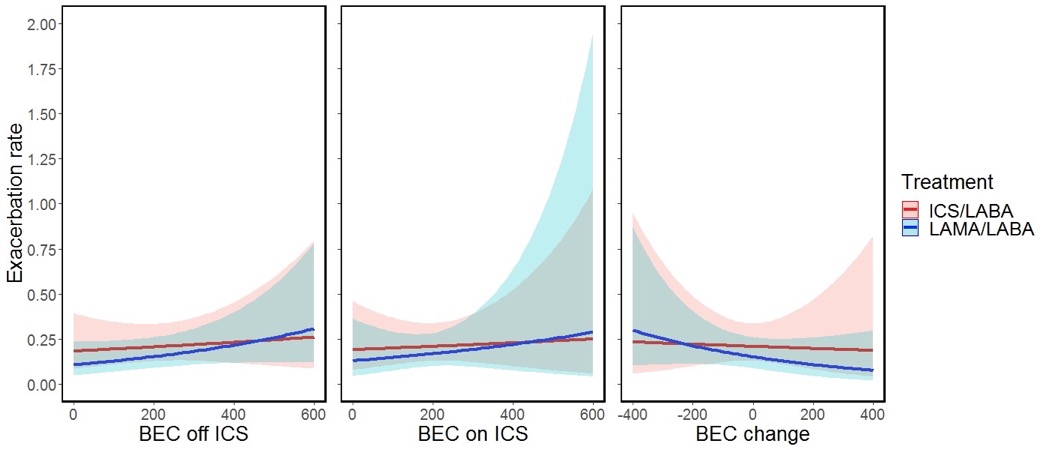

##### Figure S14. Impact of LABA/LAMA versus LABA/ICS on the frequency of **exacerbations of any severity (mild, moderate or severe)** according to (a) BEC off ICS, (b) BEC on ICS, and (c) BEC change. **Subgroup analysis only including participants that received a dose of ICS within 2 days form the BEC on ICS measurement.**

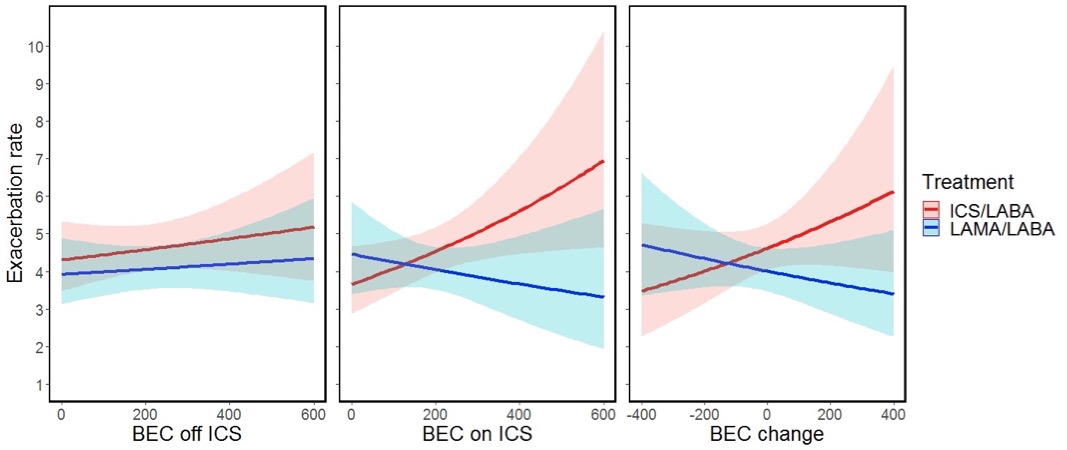

#### 2.3. Time to first exacerbation

##### Table S5: Impact of LABA/LAMA versus LABA/ICS on the time to first exacerbation. Subgroup analysis only including patients that received the last dose of their ICS within 2 days from the BEC on ICS measurement. Summary of the main results of the Cox proportional hazards analysis.

| Relevant  Parameters  Analysis | BEC | | ICS/LABA | | Three-way interaction | |
| --- | --- | --- | --- | --- | --- | --- |
|  | Coeff  (cells / mL) | p | Coeff  (Yes) | p | Coeff  (cells / mL) | p |
| **Time-to-first moderate or severe exacerbation**  BEC off ICS  BEC on ICS  BEC change | 1.220  1.640  -1.200 | **0.002**  **0.047**  **0.018** | 0.409  0.410  0.160 | 0.050  0.117  0.263 | -1.299  -1.585  1.397 | 0.056  0.142  0.128 |
| **Time-to-first severe exacerbation**  BEC off ICS  BEC on ICS  BEC change | 1.804  2.242  -1.906 | **0.025**  0.244  0.058 | 0.439  0.552  0.153 | 0.349  0.361  0.642 | -1.477  -2.504  1.134 | 0.277  0.306  0.511 |

##### Figure S15: Time-to-first **moderate or severe** **exacerbation** stratified by treatment (QVA vs SALFL) and BEC suppression by at least 170 cells/μL (-) or higher BEC change levels (+). **Subgroup analysis only including participants that received a dose of ICS within 2 days form the BEC on ICS measurement.**

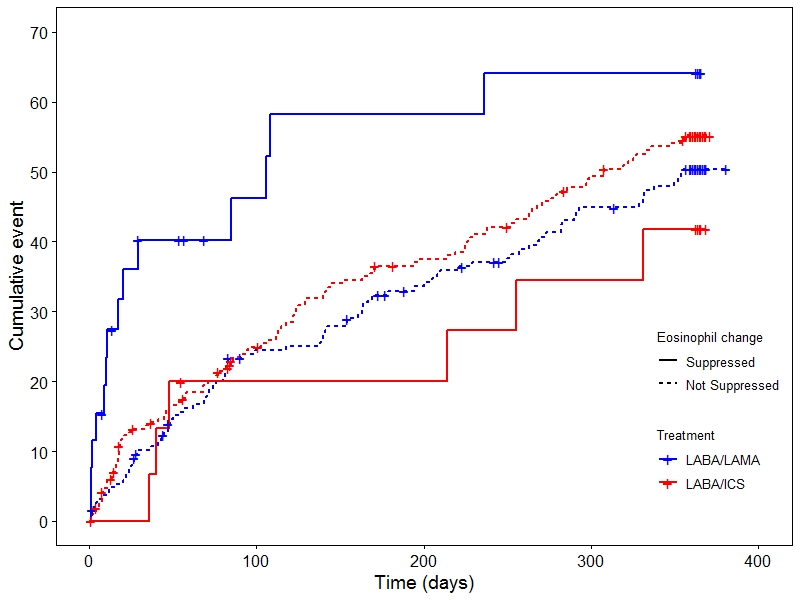

##### Figure S16: Time-to-first **severe** **exacerbation** stratified by treatment (QVA vs SALFL) and BEC suppression by at least 170 cells/μL (-) or higher BEC change levels (+). **Subgroup analysis only including participants that received a dose of ICS within 2 days form the BEC on ICS measurement.**

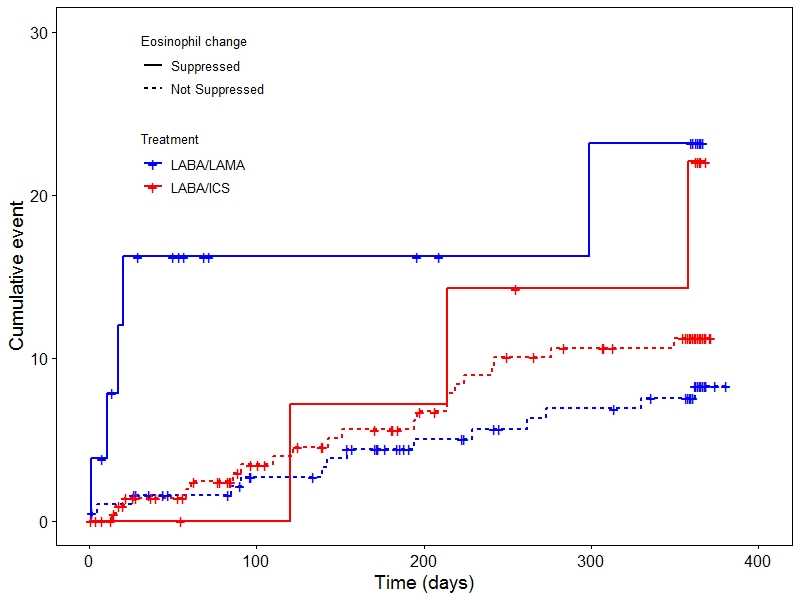

##### Figure S17: Time-to-first **exacerbation of any severity (mild, moderate or severe)** stratified by treatment (QVA vs SALFL) and BEC suppression by at least 170 cells/μL (-) or higher BEC change levels (+). **Subgroup analysis only including participants that received a dose of ICS within 2 days form the BEC on ICS measurement.**

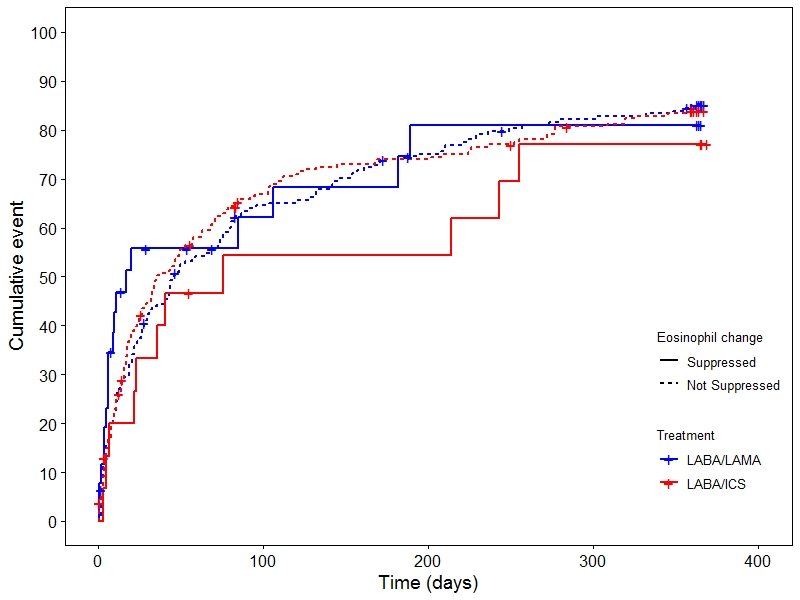

#### 2.4. Time to pneumonia

##### Table S6: Impact of LABA/LAMA versus LABA/ICS on the time to first episode of pneumonia. Subgroup analysis only including patients that received the last dose of their ICS within 2 days from the BEC on ICS measurement. Summary of the main results of the Cox proportional hazards analysis.

| Relevant  Parameters  Analysis | BEC | | ICS/LABA | | Three-way interaction | |
| --- | --- | --- | --- | --- | --- | --- |
|  | Coeff  (cells / mL) | p | Coeff  (Yes) | p | Coeff  (cells / mL) | p |
| **Time to first episode of pneumonia**  BEC off ICS  BEC on ICS  BEC change | 3.575  2.479  -4.126 | **0.006**  0.496  **0.006** | 1.380  -0.023  0.443 | 0.088  0.983  0.457 | -4.088  1.042  11.540 | 0.084  0.804  **0.002** |

##### Figure S18: Time-to-first **episode of pneumonia** stratified by treatment (QVA vs SALFL) and BEC suppression by at least 170 cells/μL (-) or higher BEC change levels (+). **Subgroup analysis only including participants that received a dose of ICS within 2 days form the BEC on ICS measurement.**

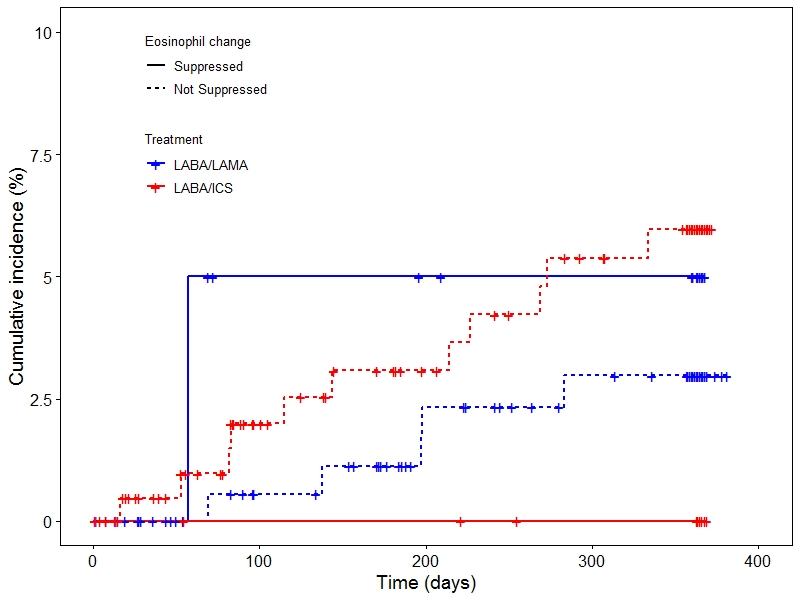

### 3. Subgroup analysis only including participants with an absolute value of BEC change of at least 200 cells/ μL.

#### 3.1. Frequency of exacerbations

##### Table S7: Impact of LABA/LAMA versus LABA/ICS on the frequency of exacerbations. **Subgroup analysis only including participants with an absolute value of BEC change of at least 200 cells/μL.** Summary of the main results of the general linear regression analysis.

| Relevant  Parameters  Analysis | BEC | | ICS/LABA | | Three-way interaction | |
| --- | --- | --- | --- | --- | --- | --- |
|  | Coeff  (cells / mL) | p | Coeff  (Yes) | p | Coeff  (cells / mL) | p |
| **Rate of moderate or severe exacerbations**  BEC off ICS  BEC on ICS  BEC change | 0.804  -1.005  -0.813 | 0.145  0.332  0.109 | 0.587  -0.545  -0.039 | 0.201  0.299  0.894 | -1.990  0.676  1.635 | **0.016**  0.686  **0.021** |
| **Rate of severe exacerbations**  BEC off ICS  BEC on ICS  BEC change | 1.185  -0.568  -1.340 | 0.383  0.823  0.296 | 0.711  -0.705  -0.301 | 0.567  0.603  0.698 | -2.958  -0.509  2.303 | 0.170  0.902  0.200 |

##### Figure S19: Impact of LABA/LAMA versus LABA/ICS on the frequency of **moderate or severe exacerbations** according to (a) BEC off ICS, (b) BEC on ICS. **Subgroup analysis only including participants with an absolute value of BEC change of at least 200 cells/μL.**

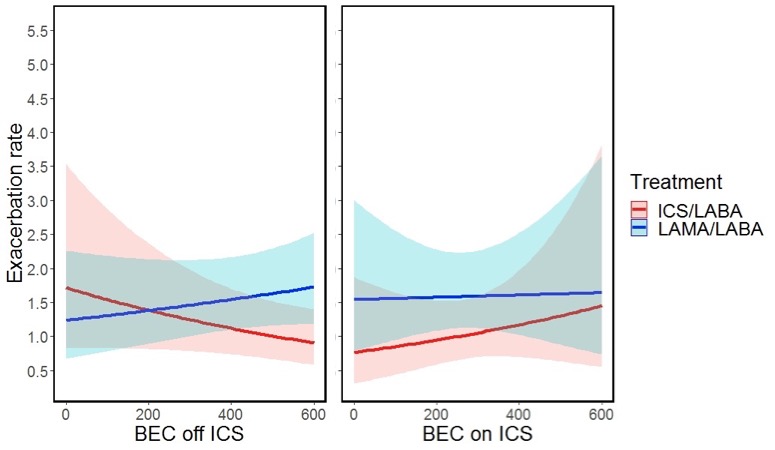

##### Figure S20: Impact of LABA/LAMA versus LABA/ICS on the frequency of **severe exacerbations** according to (a) BEC off ICS, (b) BEC on ICS. **Subgroup analysis only including participants with an absolute value of BEC change of at least 200 cells/μL.**

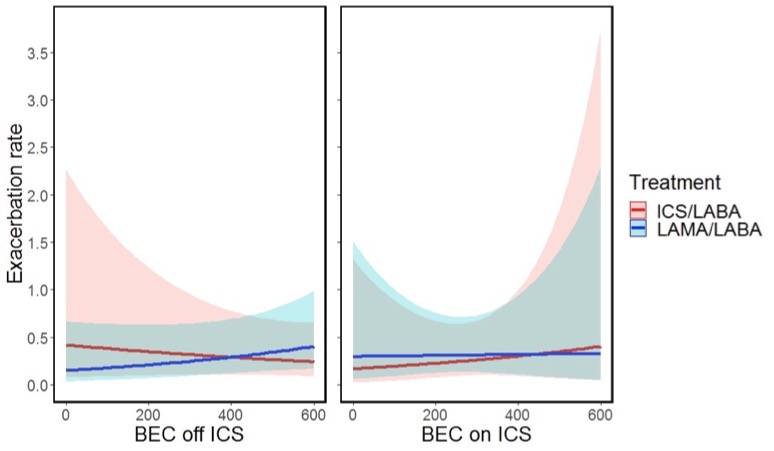

##### Figure S21: Impact of LABA/LAMA versus LABA/ICS on the frequency of **exacerbations of any severity (mild, moderate, severe)** according to (a) BEC off ICS, (b) BEC on ICS. **Subgroup analysis only including participants with an absolute value of BEC change of at least 200 cells/μL.**

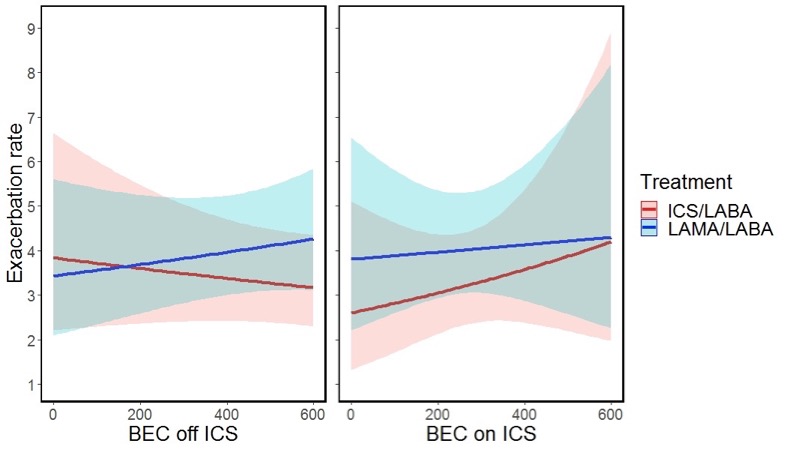

##### Figure S22: Impact of LABA/LAMA versus LABA/ICS on the frequency of **moderate or severe exacerbations** according to (a) BEC off ICS, (b) BEC on ICS, and (c) BEC change. **Subgroup analysis only including participants with an absolute value of BEC change of less than 200 cells/μL (NOT SIGNIFICANTLY CHANGED).**

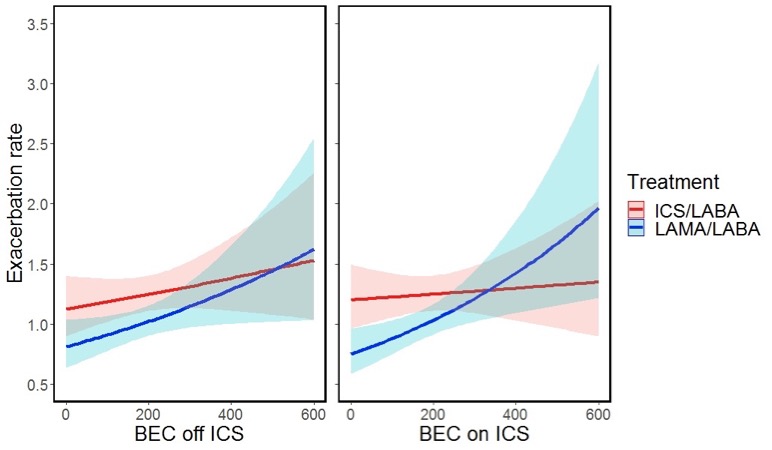

#### 3.2. Time to first exacerbation

##### Table S8: Impact of LABA/LAMA versus LABA/ICS on the time to first exacerbation. **Subgroup analysis only including participants with an absolute value of BEC change of at least 200 cells/μL.**

| Relevant  Parameters  Analysis | BEC | | ICS/LABA | | Three-way interaction | |
| --- | --- | --- | --- | --- | --- | --- |
|  | Coeff  (cells / mL) | p | Coeff  (Yes) | p | Coeff  (cells / mL) | p |
| **Time-to-first moderate or severe exacerbation**  BEC off ICS  BEC on ICS  BEC change | 0.922  -0.403  -0.917 | 0.122  0.727  0.124 | 0.688  -0.464  0.026 | 0.198  0.452  0.940 | -2.118  0.588  1.775 | **0.030**  0.757  **0.038** |
| **Time-to-first severe exacerbation**  BEC off ICS  BEC on ICS  BEC change | 2.418  0.401  -2.265 | **0.044**  0.867  0.083 | 1.515  0.071  0.614 | 0.176  0.953  0.418 | -3.019  -0.335  2.709 | 0.086  0.925  0.104 |

##### Figure S23: Time-to-first **moderate or severe exacerbation** stratified by treatment (QVA vs SALFL) and BEC suppression by at least 170 cells/μL (-) or higher BEC change levels (+). **Subgroup analysis only including participants with an absolute value of BEC change of at least 200 cells/μL.**

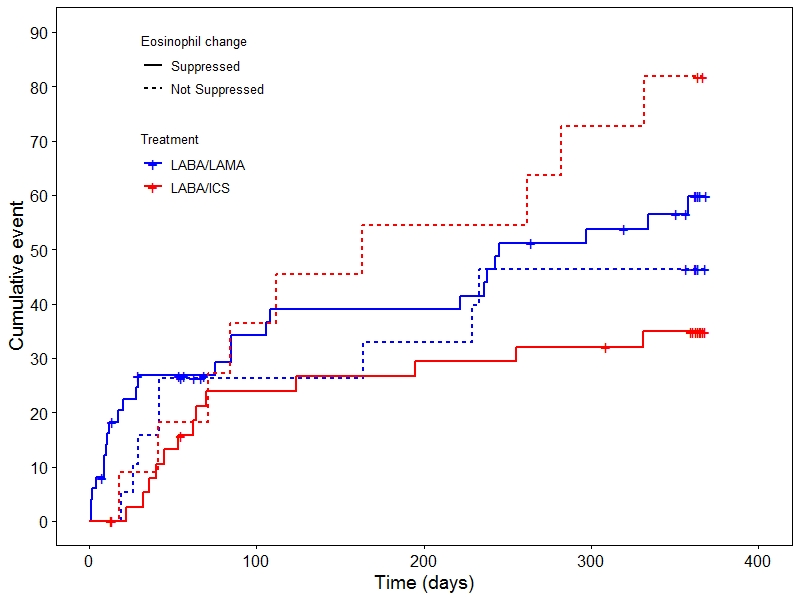

##### Figure S24: Time-to-first **severe exacerbation** stratified by treatment (QVA vs SALFL) and BEC suppression by at least 170 cells/μL (-) or higher BEC change levels (+). **Subgroup analysis only including participants with an absolute value of BEC change of at least 200 cells/μL.**

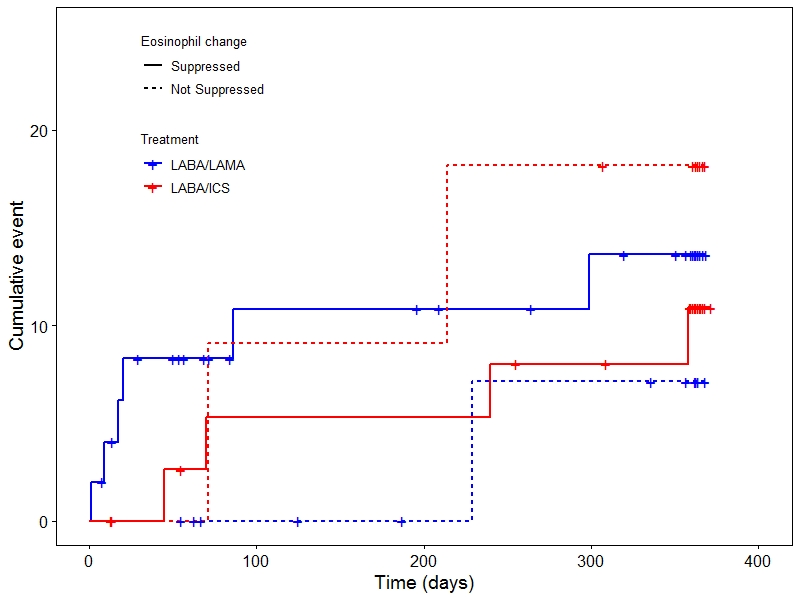

##### Figure S25: Time-to-first **exacerbation of any severity (mild, moderate, severe)** stratified by treatment (QVA vs SALFL) and BEC suppression by at least 170 cells/μL (-) or higher BEC change levels (+). **Subgroup analysis only including participants with an absolute value of BEC change of at least 200 cells/μL.**

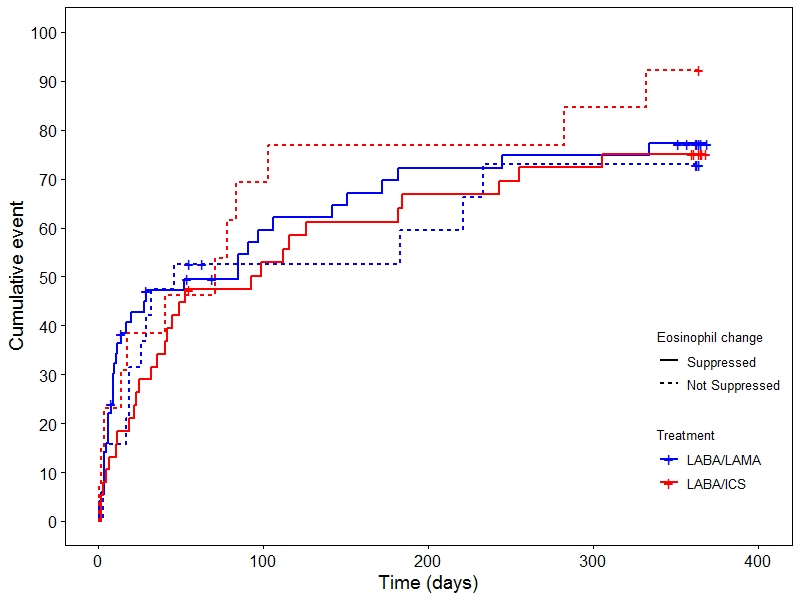

#### 3.3. Time to pneumonia

##### Table S9: Impact of LABA/LAMA versus LABA/ICS on the time to first episode of pneumonia. **Subgroup analysis only including participants with an absolute value of BEC change of at least 200 cells/μL.**

| Relevant  Parameters  Analysis | BEC | | ICS/LABA | | Three-way interaction | |
| --- | --- | --- | --- | --- | --- | --- |
|  | Coeff  (cells / mL) | p | Coeff  (Yes) | p | Coeff  (cells / mL) | p |
| **Time to first episode of pneumonia**  BEC off ICS  BEC on ICS  BEC change | 146.2  3.826  -259.6 | **<0.001**  0.602  **<0.001** | 162.1  0.998  189.3 | 0.953  0.746  0.952 | -146.8  -0.157  260.5 | **<0.001**  0.985  **<0.001** |

##### Figure S26: Time-to-first **episode of pneumonia** stratified by treatment (QVA vs SALFL) and BEC suppression by at least 170 cells/μL (-) or higher BEC change levels (+). **Subgroup analysis only including participants with an absolute value of BEC change of at least 200 cells/μL.**

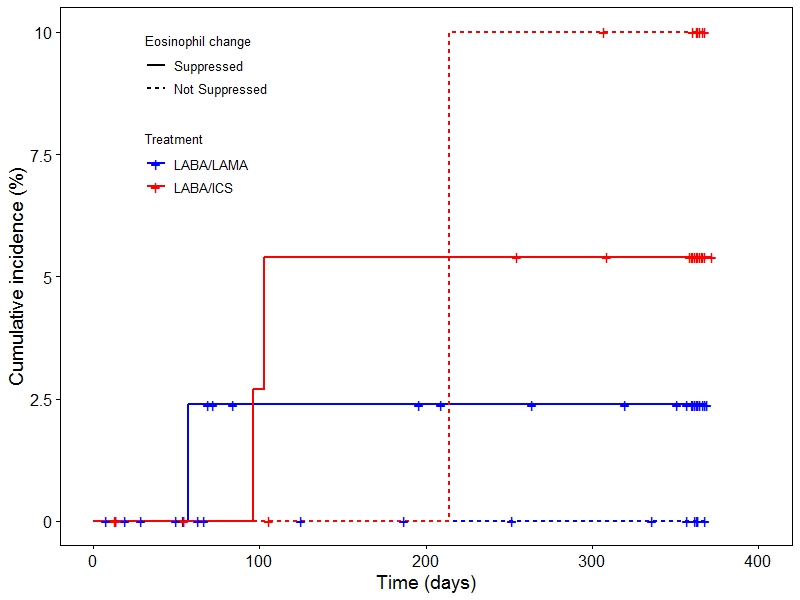

### 4. Subgroup analyses

##### Figure S27. Time-to-first **moderate or severe exacerbation.** Hazard ratio <1 favours LABA/ICS.

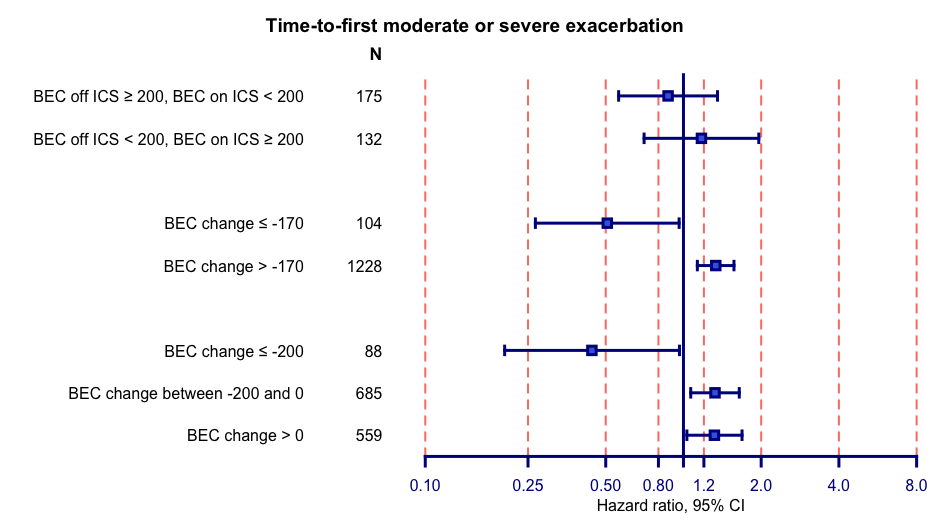

##### Figure S28. Time-to-first episode of **pneumonia** Hazard ratio <1 favours LABA/ICS.

Among patients with BEC suppression of at least 200 cells/μL, only 2 experienced pneumonia, while among those with suppression of at least 170 cells/μL, only 3 experienced pneumonia. Therefore, the corresponding HR estimates are inaccurate.

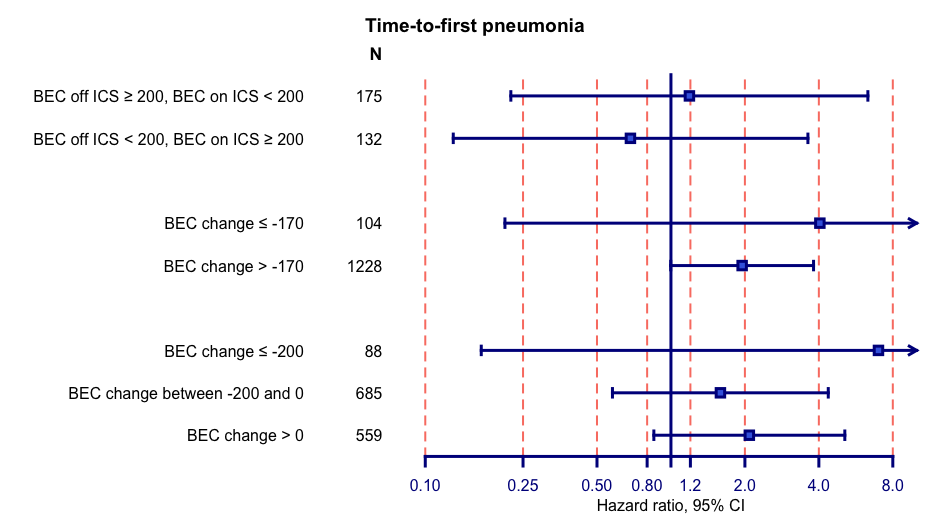

### 5. Supplementary discussion: Why have BEC on ICS signals been missed so far?

The design of RCTs could explain why the previously unidentified BEC signals on ICS were missed. First, various proportions of study participants receive ICS prior to their recruitment to RCTs resulting in initial BEC measurements reflecting a blend of BEC values both on and off ICS. Additionally, adherence to ICS therapy is frequently low in real-life where patients receive less than half of their prescribed ICS/LABA doses(1). This suggests that BEC measurements for many people that are prescribed on ICS prior to recruitment, may still represent BEC off ICS. In some trials, such as ETHOS, baseline BEC measurements are preceded by run-in periods while steroids are not allowed(2). In other trials, such as FLAME(3) or TRIBUTE(4), BEC measurements coincide with bronchodilator reversibility assessment requiring patients to temporarily halt their usual treatment. These factors could explain why the unexpected BEC on ICS signals were not previously identified. Conversely, conflicting results from retrospective studies using routinely collected data, which failed to corroborate the association between BEC and ICS treatment response, could stem from the inconsistent use of BEC measurements either on or off ICS(5, 6).

### 6. Supplementary discussion: Comparison of the findings of the FLAME and ISOLDE post-hoc analyses of BEC biomarkers.

#### 6.1. ISOLDE post-hoc analysis: Summary of findings.

The three BEC biomarkers (BEC off ICS, BEC on ICS and BEC change) were previously assessed in a post-hoc analysis of the ISOLDE, a three-year RCT comparing fluticasone propionate versus placebo in patients with COPD and an FEV_1_ of <85% predicted(7, 8). In that population, BEC change was superior in predicting treatment response to ICS compared to the other biomarkers. More specifically, ICS were proved effective in approximately 40% of patients that experienced BEC suppression during treatment with ICS, while they were not effective in cases were BEC remained largely unchanged. Importantly, BEC rose in approximately one in five patients and that was associated with a deleterious effect of ICS, characterised by 80% increase in the rate of exacerbations and more rapid FEV_1_ decline over time. Higher BEC off ICS was also predictive of decreased FEV_1_ decline over time. On the contrary, higher BEC on ICS was associated with accelerated FEV_1_ decline. Finally, neither BEC on ICS nor BEC off ICS were predictive of treatment effect on exacerbations. That was probably caused by limitations in the trial design, such as the heterogeneity of study population, that was not enriched for exacerbations, and the lack of baseline exacerbations data. Moreover, ISOLDE was conducted in the 1990s, when the standard of care was different, and the definitions of exacerbations were less standardised. Despite these limitations, our analysis demonstrated potential limitations of the use of BEC on ICS for guiding treatment decisions in COPD.

#### 6.2. Differences between FLAME and ISOLDE post-hoc analyses

The ISOLDE post-hoc analysis revealed an inverse relationship between BEC on ICS and treatment response, with higher BEC on ICS associated with accelerated FEV­_1_ decline. This inverse relationship was not confirmed in the main analysis of the FLAME study. The limitations of BEC on ICS measurements in the FLAME trial may account for this difference. As described in the main text, 66.5% of this analysis’ participants did not receive ICS for three days prior to BEC measurement, meaning that the impact of ICS on BEC had likely began to weaken by the time of the measurement. However, in sensitivity/subgroup analyses exclusive to participants showcasing significant differences between BEC off and on ICS, models based on BEC off ICS and BEC change but not those based on BEC on ICS were associated with treatment response to ICS. Moreover, a trend of higher BEC on ICS being linked to lesser treatment response to the ICS-containing regime is visible in figure 3b (rate of any exacerbations). This pattern is stronger in our sensitivity analyses (figures S13, S26), despite not reaching statistical significance. Last but not least, subgroup analyses of patients with BEC off ICS ≥200 cells/μL and BEC on ICS <200 cells/μL and vice versa and of patients with significant BEC suppression versus BEC rise also support an inverse association between BEC on ICS and treatment response to ICS, at least in a subgroup of participants.

Moreover, the two studies revealed a slightly different association between BEC change and treatment response to the ICS containing regiments. While in both studies BEC suppression and BEC during ICS treatment were associated with more and less favourable treatment response to the ICS containing regiment respectively, the intersection point differed between the two studies. More specifically, in ISOLDE the intersection point was identified at no BEC change (0 cells/μL). Therefore, a negative BEC change was associated with favourable response to ICS, while a positive BEC change was predictive of treatment failure. In FLAME, the intersection point was quantified at -170 cells/μL. This lower threshold might stem from the limitations of BEC on ICS that BEC change inherits. Moreover, LABA/ICS was compared to LABA/LAMA, a potent treatment, rather than placebo, which may require a higher ICS treatment effect to exceed the LABA/LAMA treatment effect. Both sensitivity analyses of participants showing significant BEC change and subgroup head-to-head analyses endorse the value of BEC change as a biomarker. Figure 1 displays that some patients with higher BEC off ICS, qualifying for ICS as per current guidelines, exhibit BEC rise post-ICS administration. Likewise, patients with relatively low BEC off ICS may still display BEC suppression, implying that these two biomarkers classify some patients differently. However, neither ISOLDE nor FLAME have the statistical power to compare these biomarkers, necessitating further research. Another pertinent question is whether BEC suppression or rise after ICS administration indicate distinct underlying immunological pathways.

Unlike the ISOLDE analysis, we couldn’t establish a link between BEC variables and treatment effect on pulmonary function trajectories. This could be due to the short follow-up period of one year, which might not be sufficient to evaluate treatment impact on pulmonary function decline rate. Furthermore, in ISOLDE, ICS was compared to a placebo, while FLAME compared ICS/LABA to a dual bronchodilator, known for its excellent activity on pulmonary function^(9)^.
